# A 94-bp Deletion in the Promoter of the Beta-Cell Disallowed gene *SLC16A1* causing Adult-onset Hyperinsulinism

**DOI:** 10.64898/2025.12.16.25342131

**Authors:** Jasmin J. Bennett, Hazem Ibrahim, Jonna M E Männistö, Martina Timonen, Jessica R. Hopkinson, Jonna Saarimäki-Vire, Michaelis Vasiliadis, Muhammad Faiz Muhamad, Cécile Saint-Martin, Jean-Baptiste Arnoux, Orla Neylon, Julie Okiro, Jayne A. L. Houghton, Matthew N. Wakeling, Thomas W. Laver, Matthew B. Johnson, Andrew T. Hattersley, Solja Eurola, Eliisa Vähäkangas, Hossam Montaser, Kristen Neville, Sue Mei Lau, Elizabeth Palmer, Colm Costigan, Patrick Divilly, Rachel K. Crowley, Niall Swan, James Gibney, Donal O’Shea, Yusof Rahman, Lisa G. Riley, Diego Balboa, Nick D. L. Owens, Timo Otonkoski, Sarah E. Flanagan

**Affiliations:** Clinical and Biomedical Sciences, Faculty of Health and Life Sciences, University of Exeter, Exeter, UK; Stem Cells and Metabolism Research Program, Faculty of Medicine, University of Helsinki, Helsinki, Finland; Kuopio Pediatric Research Unit (KuPRu), University of Eastern Finland, Kuopio, Finland; Robert Graves Institute of Endocrinology, Tallaght University Hospital, Dublin, Ireland; Department of Medical Genetics, AP-HP Sorbonne University, Pitié-Salpêtrière Hospital, Paris, France; Reference Center for Inherited Metabolic Diseases, Necker-Enfants-Malades University Hospital, APHP, Imagine Institute, G2M, MetabERN, Paris Cité University, 75015, Paris, France; Department of Pediatric Endocrinology, University Hospital Limerick, Dooradoyle, Co. Limerick, Ireland; Exeter Genomics Laboratory, Royal Devon University Healthcare NHS Foundation Trust, Exeter; Endocrinology, Syndney Children’s Hospital, Randwick and School of Women’s and Children’s Health, UNSW, Sydney, New South Wales, Australia; Department of Diabetes and Endocrinology, Prince of Wales Hospital, Sydney NSW 2031, Australia; School of Clinical Medicine, UNSW, Randwick, Sydney NSW 2031, Australia; School of Woman’s and Children’s Health, University of New South Wales, Sydney 2031, Australia; Centre for Clinical Genetics, Sydney Children’s Hospital, Randwick, New South Wales 2031, Australia; National Centre for Medical Genetics, Our Lady’s Hospital for Sick Children, Dublin, Ireland; Department of Endocrinology, St Vincent’s University Hospital, Dublin, Ireland; Department of Histopathology, St Vincent’s University Hospital, Dublin, Ireland; Department of Genetic Medicine, Westmead Hospital, Sydney, New South Wales 2145, Australia; Sydney Medical School, University of Sydney, Sydney, New South Wales 2006, Australia; Rare Diseases Functional Genomics, Kids Research, The Children’s Hospital at Westmead and The Children’s Medical Research Institute, Sydney, Australia; Specialty of Child and Adolescent Health, University of Sydney, Sydney, Australia; Children’s Hospital, Helsinki University Hospital and University of Helsinki, Helsinki, Finland

**Keywords:** Hyperinsulinemic hypoglycemia, Congenital hyperinsulinism, Exercise-induced hyperinsulinism, *SLC16A1*, monocarboxylate transporter 1, MCT1, Stem-cell-derived islets, Pyruvate-stimulated insulin secretion

## Abstract

Non-coding variants in the promoter region of *SLC16A1*, a beta-cell-disallowed gene encoding Monocarboxylate Transporter 1 (MCT1), cause exercise-induced hyperinsulinism (EIHI). These variants are thought to cause aberrant expression of MCT1 in pancreatic beta-cells, enabling pyruvate uptake during exercise which triggers inappropriate insulin secretion. We identified a 94-bp deletion in the *SLC16A1* promoter in 37 individuals from 11 families with childhood- or adult-onset hyperinsulinemic hypoglycemia (HI). Patient pancreatic tissue showed elevated MCT1 expression, confirming the disease mechanism. To investigate the functional impact of the deletion, we generated induced pluripotent stem cells from an affected individual and differentiated them into pancreatic stem-cell-derived islets (SC-islets). These patient-derived SC-islets showed increased *SLC16A1* gene and protein expression *in vitro* and failed to repress MCT1 expression following *in vivo* maturation in immunocompromised mice. *Ex vivo* pyruvate stimulation selectively triggered insulin secretion in variant grafts, effectively recapitulating the EIHI phenotype. In individuals with the deletion, the hypoglycemia was triggered by physical activity, carbohydrate-rich meals, and fasting, with considerable variability in age at onset and clinical severity. These findings broaden the phenotypic spectrum of *SLC16A1* promoter variants and support the inclusion of *SLC16A1* promoter analysis in the genetic evaluation of individuals with unexplained HI across all ages.

## Introduction

Hyperinsulinemic hypoglycemia (HI) is characterised by recurrent episodes of low blood glucose accompanied by inadequately suppressed insulin secretion^1^. The condition has a highly heterogeneous and age-dependent aetiology. In adults, the most frequent cause of endogenous HI is insulinoma, followed by a wide range of less common conditions^2,3^. These include endocrine, autoimmune, and genetic disorders, many of which can be difficult to clinically diagnose. Failure to identify the underlying cause can contribute to challenges in managing HI, particularly when precision therapies are available^3^.

In contrast, the underlying causes of HI presenting in early life are generally better understood and more readily diagnosed, with Congenital Hyperinsulinism (CHI) being the most common form. CHI is a monogenic disorder caused by pathogenic variants in several genes encoding key components of the insulin secretion pathway in pancreatic beta-cells^1,4^. Although rare, some case reports have described adult-onset HI due to pathogenic variants in the known CHI genes. In most of these adult cases the genetic diagnosis was made following the identification of the variant in a child with HI from the same family^5–8^.

*SLC16A1* is routinely included in the list of genes for CHI, although some laboratories do not include the non-coding promoter region on their HI gene panels^3,4^. The gene encodes monocarboxylate transporter 1 (MCT1), which facilitates the transport of pyruvate and other monocarboxylates across cell membranes. While MCT1 is widely expressed in most tissues, its expression is actively repressed (or disallowed) in pancreatic beta-cells^9^.

Pathogenic non-coding variants in the *SLC16A1* promoter region have been reported in 13 individuals from four families. These individuals were diagnosed with HI at various ages, ranging from infancy to late adolescence^10,11^. Clinical studies demonstrated that pyruvate and anaerobic exercise triggered inappropriate insulin secretion leading to hypoglycemia in these patients^12–14^. These findings led to the hypothesis that the promoter variants result in aberrant expression of MCT1 in pancreatic beta-cells. This enables the uptake of pyruvate into the beta-cells following its release into the blood stream during exercise causing beta-cell membrane depolarisation which triggers insulin secretion, even under hypoglycemic conditions^10^.

Further support for this proposed mechanism came from transgenic mouse models, in which doxycycline-induced beta-cell–specific overexpression of MCT1 caused isolated islets to secrete insulin in response to pyruvate. Furthermore, these mice also secreted insulin in response to exercise, recapitulating the human phenotype^15^. No studies to date have directly assessed MCT1 expression in pancreatic tissue from humans with *SLC16A1*-HI.

Recently, non-coding variants affecting a second beta-cell-disallowed gene, *HK1*, have been established as a cause of CHI^16,17^. These variants disrupt a *cis*-regulatory element critical for maintaining the repression of hexokinase 1 within the beta-cell. Aberrant expression of HK1 is hypothesised to lower the glucose threshold for insulin secretion, leading to inappropriate insulin release during hypoglycemia^16^. The discovery of variants in *HK1* and *SLC16A1* highlight the critical role of non-coding regulatory variants in the etiology of CHI, whereby inappropriate expression of a normally repressed, yet functionally intact, protein in beta-cells drives dysregulated insulin secretion.

In this study, we report a novel 94-bp deletion in the promoter region of *SLC16A1,* identified in 37 individuals from 11 families with HI presenting across a wide range of ages. To investigate the pathophysiological impact of this variant, we analysed patient pancreatic tissue and developed a human stem-cell-based model that enabled functional studies. These findings expand the clinical understanding of HI and highlight the importance of screening the *SLC16A1* promoter region in all individuals, including adults, with persistent hyperinsulinemic hypoglycemia of unknown cause.

## Methods

### Index family

The proband was clinically diagnosed with HI in adulthood following repeated episodes of light-headedness and fainting that had persisted since adolescence but had increased with frequency and severity with age. They were prescribed with diazoxide (9 mg/kg/d), but this did not reduce the severity or frequency of hypoglycemic episodes. They subsequently underwent a distal pancreatectomy. Histological analysis of the resected tissue revealed no evidence of a neuroendocrine neoplasm. Post-surgery, their glucose levels normalised. However, HI returned and persisted within months (blood glucose 2.4 mmol/L, insulin 194 pmol/L, C-peptide 3.52 mcg/L, HbA1c 27 mmol/mol).

They had a family history of adult-onset hypoglycemia, which affected a parent, sibling, and two cousins. Their affected parent was diagnosed with hypoglycemia in late adulthood (blood glucose 2.9 mmol/L, insulin 174 pmol/L). They were reported to be intolerant to diazoxide and underwent a partial pancreatectomy. Post-surgery their blood glucose levels remained within the normal range.

The proband’s sibling was diagnosed with HI in adolescence (glucose 2.3 mmol/L, insulin 152 pmol/L). In adulthood, multiple imaging studies (endoscopic ultrasound, CT pancreas, gallium DOTA-TOC PET/CT, MRI pancreas and ^68^Ga-DOTA-extendin-4 PET/CT) had failed to detect an insulinoma or a localised lesion of hyperplasia within the pancreas. They underwent a partial pancreatectomy due to a partial response to diazoxide and a poor response to octreotide, which worsened the symptoms. They continue to experience recurrent hypoglycemic episodes which are not medically treated.

Two of the proband’s cousins were clinically diagnosed with HI, one in adolescence and the other at in adulthood. The cousin diagnosed in adulthood reported experiencing symptoms consistent with hypoglycemia since childhood which had not been therapeutically managed. The cousin diagnosed in adolescence had undergone unsuccessful treatment trials with diazoxide and lanreotide. They continue to experience recurrent hypoglycemic episodes which are not medically treated.

All five affected individuals in the index family report significant disruption to their daily lives due to frequent hypoglycemic episodes. They are attempting to manage their condition through dietary strategies, including eating small, frequent meals with a low glycemic index every 2.5 hours, and carbohydrate loading before and after exercise.

### Molecular genetic testing

Screening of the *MEN1* and *CDKN1B* genes was performed on leukocyte DNA from the female proband to investigate the possibility of endocrine neoplasia. No pathogenic variants were detected. Two years later, the proband reached out to the Exeter laboratory, requesting to be included in their ongoing research studies focussed on genetic discovery in CHI. In consultation with the family’s clinical team, leukocyte samples were collected from proband and their affected sibling, parent, and two cousins for DNA extraction and whole genome sequencing (WGS).

WGS was performed to a mean read depth of 32 using the BGISeq-500 platform on samples from all five individuals. The sequence data was aligned using BWA MEM 0.7.15 and processed with a pipeline based on the GATK best practices (Picard version 2.7.1, GATK version 3.7). Variants were annotated using Alamut Batch Standalone version 1.11 (Rouen, France).

Variants present in gnomAD v4.1 were filtered out. The remaining novel variants were annotated using snATAC-seq signal peaks in beta_1 cells, downloaded from the publication’s repository (https://catlas.org/catlas/index.php) to indicate those in regions of open chromatin in beta-cells^18,19^.

### Replication cohort

Replication studies were performed by targeted next-generation sequencing (tNGS) using an international cohort of 1,258 probands who had been referred to the Exeter Genomics Laboratory for CHI testing (median age at diagnosis of HI: 2 weeks [IQR: birth–8 months]) with no disease-causing variants identified^20^. Two further probands with the 94-bp deletion were identified by the Department of Medical Genetics, Pitié-Salpêtrière Hospital (Paris, France) and Westmead Hospital (Sydney, Australia) following tNGS of the known CHI genes (**Supplementary Methods**). The case from Australia has been recently published^21^.

When a pathogenic variant was identified, samples from family members were tested for the variant by tNGS or by Sanger sequencing. Sanger sequencing involved PCR amplification of the region of interest (primer sequences are listed in **Supplementary Table 1**). PCR products were sequenced on an ABI3730 capillary sequencer (Applied Biosystems, Warrington, UK) and analysed using Mutation Surveyor v3.24 software (SoftGenetics, State College, PA, USA) or SeqScape (Thermo Fisher Scientific, Waltham, Massachusetts, USA).

### Haplotype analysis

SNP array genotyping (Erasmus MC, Rotterdam, Netherlands) was performed on DNA of 9 unrelated individuals using Illumina GSA Beadchip GSA MD (Illumina GSA Arrays ‘Infinium iSelect 24x1 HTS Custom Beadchip Kit’). In-house bioinformatics pipelines were developed to search for a shared haplotype across the promoter region of *SLC16A1*. A haplotype was considered shared when two or more individuals shared 300 or more consecutive variants.

### Immunohistochemistry studies

Formalin-fixed paraffin-embedded pancreatic tissue was available for analysis from the proband, sibling and parent of the index family and a second proband (**Figure 2 and Supplementary Figure 1**)). Immunohistochemistry, using Akoya Biosciences OPAL 6-Plex detection kits (NEL861001KT), was performed on 4-μm-thick tissue sections, and slides were scanned at x40 using the Phenolmager HT 2.0 whole slide scanner (Akoya Biosciences). For comparison, pancreatic tissue from individuals with *HK1*-HI and non-HI controls, age matched to the cases, were also examined. Quantitative image analysis was carried out in QuPath (V0.4.3)^22^. Initially, a pixel classifier was developed to create an annotation layer for tissue, then within that annotation layer, the islets were classified using a random trees classifier. Cells within the islets were segmented using the Stardist plugin^23^, and then classified based on hormone and MCT1 expression. The median fluorescence intensity (MFI) of MCT1 in all insulin positive cells was plotted to determine the MCT1 staining intensity in beta-cells. All images were manually checked to remove any areas of misclassified islets, tissue folds included in analysis and islets that bordered the edge of the tissue to reduce “edge effect”. Additional details are provided in the **Supplementary Methods.**

### Epigenomic analysis

To assess chromatin accessibility at the *SLC16A1* promoter across 222 cell types, we downloaded processed signal tracks in bigwig format from the publication’s associated repository (https://catlas.org/catlas/index.php)^18^. For beta-cells we visualised the beta_1 cluster as defined^19^, as a beta-cell cluster exhibiting robust chromatin accessibility over *INS*. To contrast to other cell types, we ordered the heatmap by maximal chromatin accessibility (**Supplementary Figure 2**).

### Generation of a patient-derived induced pluripotent stem cell model and in vitro endocrine differentiation

Dermal fibroblasts were obtained from a skin biopsy of an adult male (**Supplementary Figure 3**) with a heterozygous 94-bp deletion in the *SLC16A1* promoter. The fibroblasts were reprogrammed using CRISPR-activator system as previously described and summarised in **Supplementary Methods**^24^. The formed HEL400 iPSC lines were verified using Sanger sequencing to confirm the deletion (**Supplementary Figure 3B, C**) and tested negative for mycoplasma contamination. The iPSC line HEL24.3, derived from human neonatal foreskin fibroblasts, which is wild type for the *SLC16A1* promoter, was used as a control. Generated stem cell lines were cultured on Matrigel-coated plates with E8 medium and passaged using 0.5 mmol/L EDTA (Life Technologies, 15575–038; USA) in PBS as a dissociation agent. Two iPSC clones (HEL400.6 and HEL400.7) derived from the same patient reprogramming experiment were characterized and are referred to as *SLC16A1* variant (Var; HEL400.6 Var1 and HEL400.7 Var2), while the control line HEL24.3 is referred to as wild type (WT).

The iPSCs were differentiated towards the pancreatic lineage to generate pancreatic endocrine cells using our previously published protocol with minor modifications^25,26^. Flow cytometry was performed to quantify the percentage of definitive endoderm-positive cells at stage 1. Immunohistochemistry confirmed flow cytometry findings. Details are provided in the **Supplementary Methods.**

### Static glucose- and pyruvate stimulated insulin secretion (GSIS and PSIS) assays

Static GSIS and PSIS assays were performed on cells that had been sequentially treated with 2.8 mmol/L glucose, followed by 16.8 mmol/L glucose for GSIS or 0.5 mmol/L pyruvate for PSIS, and final depolarization with 30 mmol/L KCl. Insulin content results were normalised to the DNA content of the beta-cell fraction, calculated from flow cytometry insulin-positive cell percentage.

### In vivo animal implantation studies

Implantations were performed on randomised 4- to 8-month-old male mice. Briefly, stage 7 WT and Var SC-islets equivalent to 1.5 million cells (600 clusters) were loaded in PE-50 tubing and implanted under the kidney capsule. Mouse serum samples were collected monthly from the saphenous vein and stored at −80°C for human C-peptide analysis. Blood glucose levels were measured using ContourXT and ContourNext strips and human-specific plasma C-peptide was measured with the Ultrasensitive C-peptide ELISA kit (Mercodia).

### Ethical considerations

Informed consent was obtained from all participants reported in this study. The genetic studies performed in Exeter were approved by the Wales Research Ethics Committee 5 through the Genetic Beta-Cell Research Bank (22/WA/0268) and were conducted in accordance with the principles of the Declaration of Helsinki. The iPSCs were generated and used with informed consent from the donor according to the approval of the coordinating ethics committee of the Children’s Hospital at Westmead (2019/ETH11736) and the Helsinki and Uusimaa Hospital District (no. 423/13/03/00/08). The Animal care and experiments were conducted in Biomedicum Helsinki animal facility as approved by the National Animal Experiment Board in Finland (ESAVI/47888/2023).

## Results

### Detection of an SLC16A1 94-bp deletion in 37 individuals

WGS was performed on five affected members of the index family. WGS revealed 1,062 heterozygous variants present in all five individuals but absent from gnomAD v4.1^27^. All these variants were non-coding. Twenty-three of the variants mapped to regions of open chromatin in pancreatic beta-cells, suggesting potential regulatory effects on gene expression in this cell type (**Supplementary Table 4**). One of these variants was a novel heterozygous 94-bp deletion, GRCh38:1:112,956,297-112,956,390del, within the *SLC16A1* promoter region (**Figure 1**). This variant was prioritised for further study because it is situated just 7-bp from the 25-bp insertion identified in family 2, and 88-bp from the single nucleotide variant reported in family 1 by Otonkoski *et al* (**Figure 1**)^10,12^.

**Figure 1.**
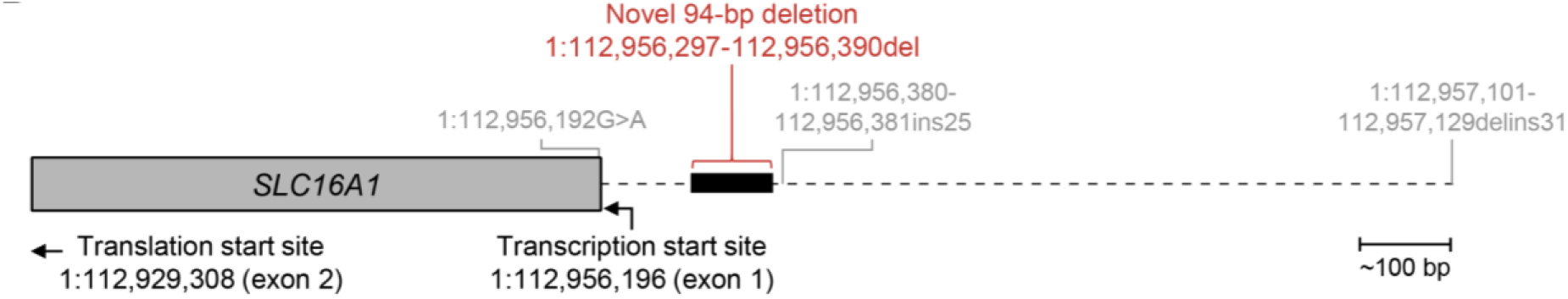
Genomic location of the *SLC16A1* 94-bp promoter deletion. Genomic coordinates are based on GRCh38, with *SLC16A1* encoded on the minus strand. The variant was called as: GRCh38:1:112,956,279CCGCCCCCTCCCCGCCACACAGACATCCGAACTGCAGCCCGCGCCTCCACACGCTTTCAGCCGCGCGCGCCCTCTAGCTCGCCCGCGCGCGCCGG>C in VCF. The variant is described as GRCh38:1:112,956,297-112,956,390del according to HGVS. The difference in nomenclature between the HGVS description and VCF call is due to a 17-bp repeated motif at the beginning and end of the deleted region. The deletion is indicated by a black box relative to the transcription start site. Previously reported variants by Otonkoski *et al.* are shown in grey ^10^. Created in BioRender. Bennett, J. (2026) https://BioRender.com/2bntfnx.

Replication studies identified the same heterozygous *SLC16A1* 94-bp deletion in ten additional probands. Co-segregation analysis confirmed the presence of this variant in 20 affected relatives across six of these families. In one proband the 94-bp deletion had arisen *de novo*. Two clinically affected obligate heterozygotes in two families were also identified, including one in the index family. In total, the 94-bp deletion was identified in 37 individuals, comprising 11 probands and 26 family members.

### Variant classification

Based on the strong genetic evidence and the demonstration of aberrant MCT1 expression in patient pancreatic tissue (results below), the 94-bp deletion (GRCh38:1:112,956,297-112,956,390del) was classified as ‘pathogenic’ according to established variant interpretation guidelines (**Supplementary Table 5**)^28–30^.

### Founder variant in the Irish population

As eight of the 11 probands (73%) reported Irish ancestry, we performed SNP array analysis to investigate the possibility of a founder variant. This analysis included one individual from each of the eight Irish families and one proband referred from Australia. A shared haplotype spanning at least ~1.9 Mb across the *SLC16A1* gene was identified in all individuals of Irish ancestry, consistent with a founder variant in this population (**Supplementary Figure 4**). The absence of this haplotype in the non-Irish proband, combined with the identification of a *de novo* variant in one individual, confirms that this 94-bp deletion is a recurrent variant that has arisen independently on multiple alleles over time.

### Clinical characteristics of the 37 individuals with the SLC16A1 variant

The clinical features of the 37 individuals with the *SLC16A1* variant are shown in **Table 1**. At diagnosis the median blood glucose levels were 2.4 mmol/L with a paired insulin of 37 pmol/L.

**Table 1:** Summary of clinical features for 37 individuals heterozygous for the 94-bp *SLC16A1* promoter deletion.

|  | Summary data |
| --- | --- |
| Clinical diagnosis of HI % (n) | 62 (23/37) |
| Anecdotal hypoglycemia symptoms % (n) | 38 (14/37) |
| Female sex % (n) | 57 (21/37) |
| Median age at last follow up in years [IQR] (n) | 46 [36-64] (33) |
| Median age at diagnosis in years [IQR] (n) | 40 [29-49] (27) |
| Childhood onset (<10 years) % (n) | 4 (1/27) |
| Adolescence onset (10-18 years) % (n) | 19 (5/27) |
| Adult-onset (>18 years) % (n) | 78 (21/27) |
| Median blood glucose at diagnosis mmol/L [IQR] (n) | 2.4 [2.2-2.8] (20) |
| Median paired insulin at diagnosis pmol/L [IQR] (n) | 37 [35-127] (14) |
| Median HbA1c mmol/mol [IQR] (n) | 28 [24-29] (13) |
| Evidence of exercise trigger % (n) | 51 (19/37) |
| Confirmed on formal exercise stress test % (n) | 16 (3/19) |
| Anecdotal exercise-related symptoms % (n) | 84 (16/19) |
| Evidence of fasting trigger % (n) | 38 (14/37) |
| Confirmed on formal fasting test % (n) | 86 (12/14) |
| Anecdotal fasting-related symptoms % (n) | 14 (2/14) |
| Additional triggers noted (n) | High carbohydrate load (3)<br>Protein-rich meals (1)<br>Emotional stress (3)<br>Alcohol (4) |
| History of seizure % (n) | 16 (6/37) |
| Treatment for HI % (n) | 49 (18/37) |
| Surgical treatment hypoglycemia resolved % (n) | 11 (2/18) |
| Surgical treatment hypoglycemia persisted % (n) | 17 (3/18) |
| Medical (diazoxide) treatment % (n) | 72 (13/18) |

The number of individuals (n) for whom data was available is shown in parentheses. HI: hyperinsulinemic hypoglycemia; IQR: interquartile range

The median age at diagnosis of hypoglycemia was 40 years **(Table 1)**. Most, 21 (78%), were diagnosed in adulthood (>18 years), 5 (19%) in adolescence (10–18 years) and one (4%) individual during early childhood (1-2 years). Notably, at least 12 adults diagnosed between ages 30 and 78 years reported experiencing hypoglycaemic symptoms for many years before their clinical diagnosis, some dating back to childhood. Interestingly, two family members were reported to be asymptomatic at the time of genetic testing, but subsequent clinical investigations confirmed hypoglycemia.

Treatment was documented in 18 individuals **(Table 1)**. Five underwent partial distal pancreatectomy between 33 and 72 years of age, including three individuals from the index family. Three of the five individuals continued to experience hypoglycemia post-operatively. Thirteen individuals who had not undergone pancreatectomy received diazoxide therapy; this was continued in all but one case who was reported to be non-responsive. The individual with childhood-onset HI was treated with a high dose diazoxide (15 mg/kg/day). In one individual a trial with octreotide was discontinued due to reported worsening of hypoglycemic symptoms.

A low HbA1c, with a median of 28 mmol/mol [IQR: 24-29] was a notable feature, suggesting that at least modest hypoglycemia is present even in the absence of symptoms. In 16% (6/37) of individuals, a history of seizures indicated that hypoglycemia was sufficiently severe to cause severe neuroglycopenia.

### Triggers for hypoglycemia in individuals with SLC16A1-HI

Individuals reported multiple triggers for hypoglycemia. Nineteen individuals reported symptomatic hypoglycemia triggered by physical activity (**Table 1**). In 3/19 this was confirmed by formal exercise stress testing. Fourteen individuals, including 8 who had identified exercise as a trigger, reported prolonged fasting was a likely precipitant based on characteristic episodes. This was confirmed on a formal prolonged fasting test in 12 individuals (**Supplementary Table 6**). Individuals also reported hypoglycemia symptoms triggered by a high carbohydrate load (*n*=3), protein-rich meals (n=1), emotional stress (*n*=3), or alcohol consumption (*n*=4).

### Immunohistochemistry demonstrates MCT1 presence in patient pancreatic beta-cells

Pancreatic tissue from four individuals who had undergone partial pancreatectomy was analysed by immunohistochemistry. MCT1 staining colocalised with insulin, confirming its presence within the pancreatic beta-cells of affected patients, but not in age-matched controls (**Figure 2A and Supplementary Figure 1**). MCT1 expression was specifically localised to the beta-cell membrane, consistent with its known role as a membrane-bound transporter. Quantitative analysis demonstrated a significantly higher median fluorescence intensity (MFI) of MCT1 in islets from individuals with the *SLC16A1* promoter deletion compared to controls (p ≤ 0.0001; **Figure 2B**).

**Figure 2.**
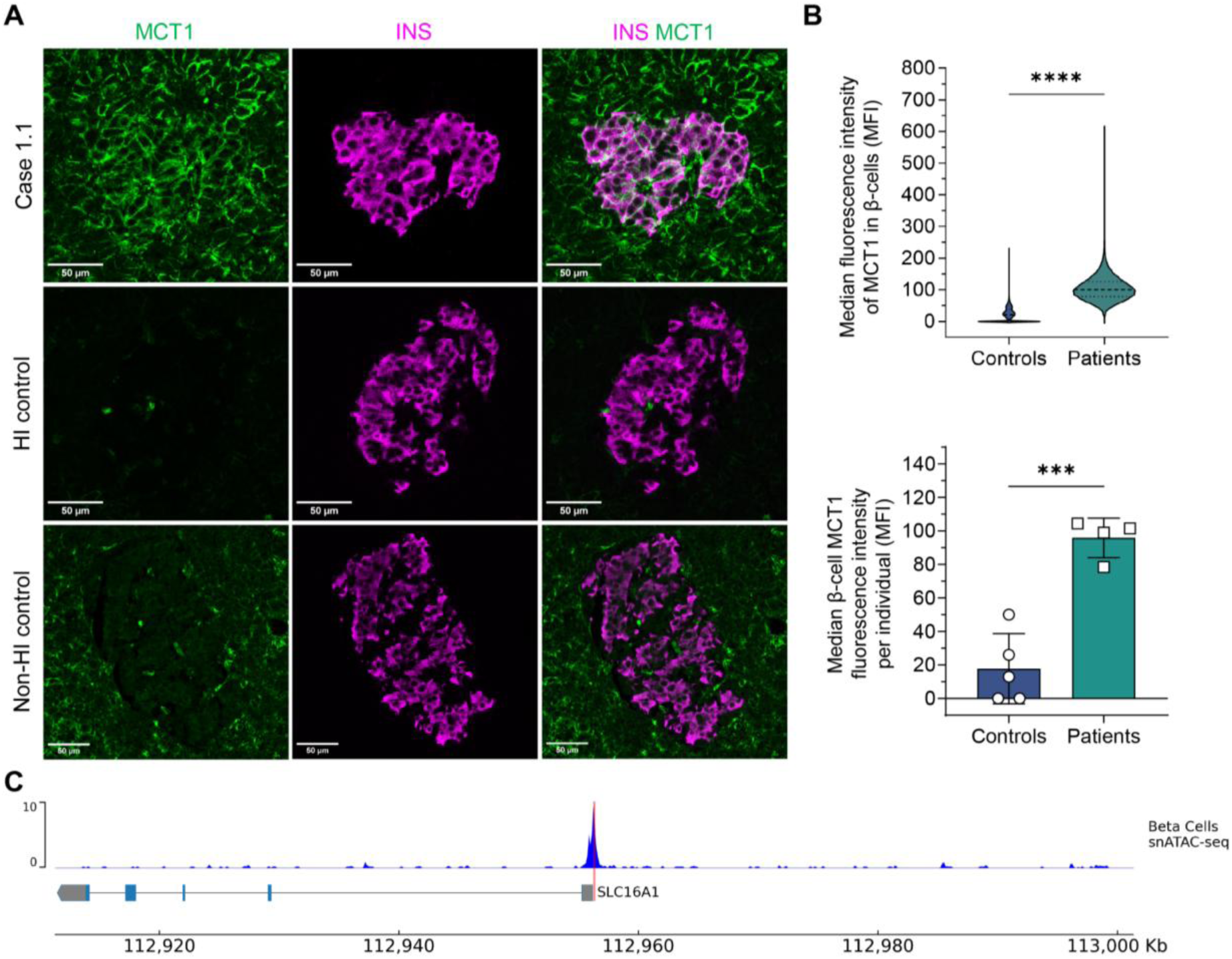
MCT1 expression in pancreatic tissue with *SLC16A1* promoter deletion and chromatin accessibility in human beta-cells. (A) Confocal imaging of MCT1 (green) and Insulin (magenta) in pancreatic tissue from the index proband with the 94-bp *SLC16A1* promoter deletion demonstrate that MCT1 is expressed in the exocrine acinar cells and endocrine beta-cells in patients with the *SLC16A1* 94-bp deletion but absent in pancreatic beta-cells of hyperinsulinism (HI) control and non-HI control donors. Scale bar: 50 µm. Images from additional cases are shown in Supplementary Figure 1. (B) Top panel: violin plot displaying median fluorescence intensity (MFI) of MCT1 in beta-cell membranes from Controls (HI and non-HI; *n*=5 patients, 97,978 beta-cells) and patients with the *SLC16A1* 94-bp deletion (*n*=4 patients, 107,219 beta-cells). Bottom panel: bar plot showing median MFI per individual, presented as means ±SD. Statistical significance was measured using unpaired *t*-test; \*\*\**p*<0.001, \*\*\*\**p*<0.0001. (C) snATAC-seq data in human beta-cells showing chromatin accessibility centred on the *SLC16A1* promoter region (GRCh38:1:112,956,297-112,956,390del) in normalised signal per million reads. The red highlighted segment indicates the 94-bp deletion region, coinciding with the peak of chromatin accessibility in beta-cells.

### Epigenomic results

We investigated epigenomic signals spanning the *SLC16A1* variant region. First, we confirmed that *SLC16A1* is broadly expressed across adult tissues, with notable repression in pancreas (**Supplementary Figure 5A**), and that its expression decreases progressively over stem-cell pancreatic differentiation (**Supplementary Figure 5B**)^31^. Single-cell atlas chromatin accessibility data from 222 fetal and adult cell types^18^, including pancreatic beta-cells^19^, revealed a distinct peak of chromatin accessibility overlapping the 94-bp deletion (**Figure 2C**). We observed that cell types expressing *SLC16A1* exhibit far greater promoter chromatin accessibility compared to pancreatic beta-cells (**Supplementary Figure 2**), consistent with active transcriptional machinery in expressing cells. These findings imply that as-yet unidentified factors bind at the 94-bp deleted region to confer repression, producing a peak of chromatin accessibility that is markedly smaller than *SLC16A1*-expressing cell types.

### Patient-derived SC-islets exhibit upregulated SLC16A1 expression

Patient-specific induced pluripotent stem cells (iPSCs) were derived from patient fibroblasts. MCT1 and *SLC16A1* expression levels in patient fibroblasts were comparable to those in control fibroblasts from four healthy donors (**Supplementary Figure 3**).

The patient iPSC clones alongside the healthy control iPSCs were next differentiated into the pancreatic endocrine lineage (**Figure 3A**)^24,25^, with additional quality control data provided in the supplementary results (**Supplementary Figures 6, 7**). Both wild type (WT) and *SLC16A1* variant (Var) lines produced endocrine cells with sufficient INS⁺ (insulin-positive) beta-cells (**Figure 3B**). During differentiation, we observed a progressive decline in *SLC16A1* expression in both WT and Var lines, with expression diverging after endocrine cells have formed leading to 6-fold higher expression in Var cells at week 7 of the final differentiation stage 7 (S7w7) (**Figure 3C**). Immunohistochemistry demonstrated membranous MCT1 staining in all cells at S7w3, and more prominent in Var SC-islets than in WT at S7w7 (**Figure 3D**). Quantification of MCT1-positive area within the E-cadherin-defined beta-cell membrane confirmed significantly higher MCT1 levels in Var compared to WT at S7w7 (**Figure 3E**).

**Figure 3.**
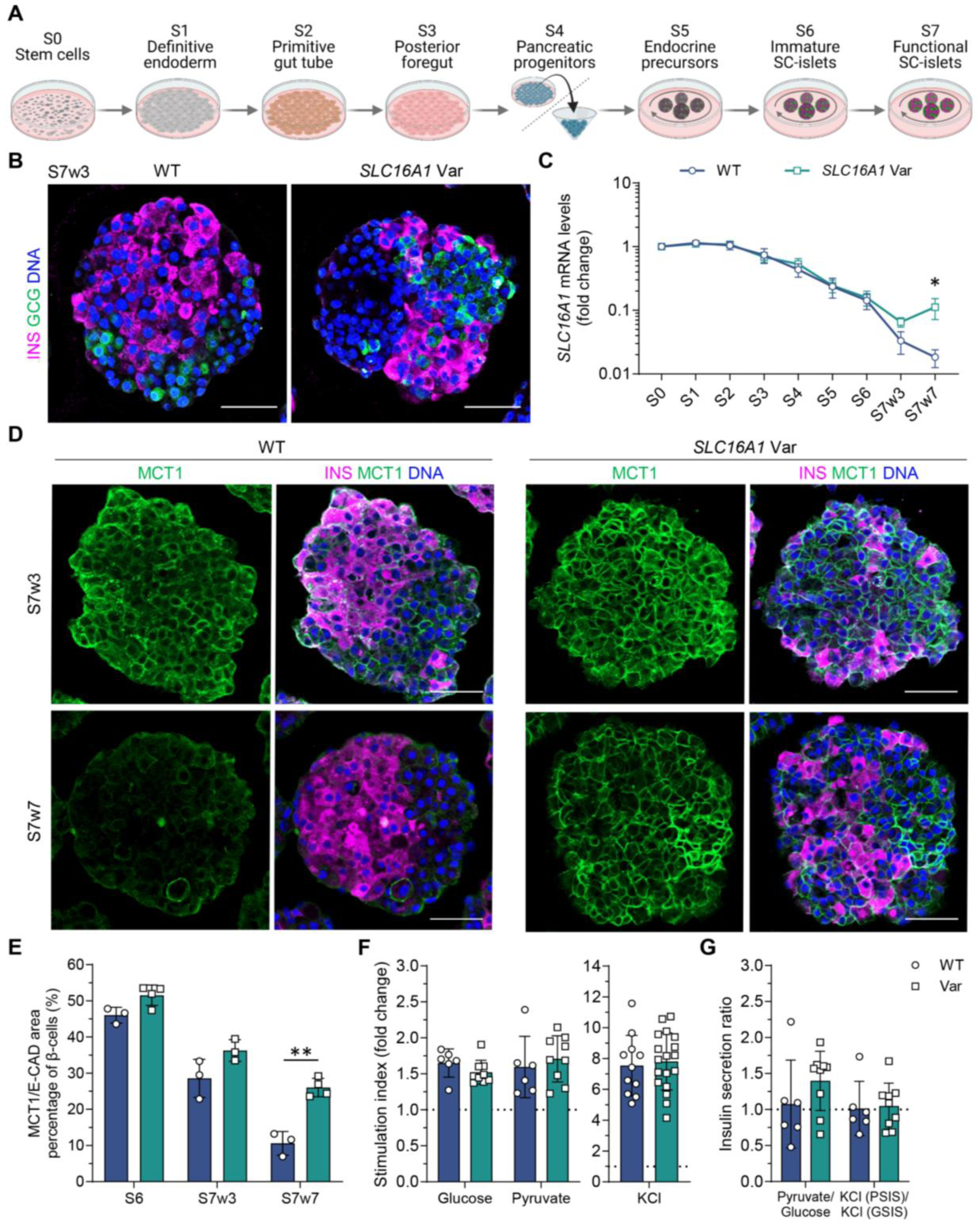
Upregulated *SLC16A1* expression in the variant SC-islets. (A) Schematic representation of the differentiation stages and formats of iPSCs towards SC-islets. Illustration made with BioRender.com. (B) Immunofluorescence images showing insulin-positive (INS^+^) and glucagon-positive (GCG^+^) cells for SC-islets at S7 week 3 (S7w3). Scale bar, 50 µm. (C) Relative gene expression levels of *SLC16A1* between control and variant cell lines across the differentiation stages (*n*=3–8). (D) Immunofluorescence images showing INS^+^ and MCT1^+^ cells at S7w3 and S7w7. Scale bar, 50 µm. (E) Quantification of the percentage of MCT1 signal area localised within the E-cadherin (E-CAD)-defined membrane region of INS⁺ cells (*n*=3–4). (F) Insulin secretion stimulation index from static GSIS and PSIS at S7w7, shown as fold change relative to 2.8 mmol/L glucose. Conditions: Glucose (16.8 mmol/L), Pyruvate (0.5 mmol/L), and KCl (30 mmol/L) (*n*=6–9). (G) Insulin secretion ratio between pyruvate and glucose stimulation and between KCl-response in PSIS versus GSIS at S7w7 (*n*= 6–9). Statistical significance in (C, E–G) was measured using unpaired *t*-test. Data are presented as means ±SD, \**p*<0.05, \*\**p*<0.01.

To assess whether elevated *SLC16A1* expression in Var cells enhances pyruvate-stimulated insulin secretion, we performed static glucose- and pyruvate-stimulated insulin secretion (GSIS and PSIS) assays at S7w7. Both WT and Var cells exhibited robust insulin secretion in response to glucose, pyruvate, and KCl, with comparable pyruvate stimulation index despite lower *SLC16A1* expression in WT cells (**Figure 3F**), suggesting that residual MCT1 expression may suffice to mediate pyruvate uptake. However, Var cells showed a trend of increased pyruvate-stimulated insulin secretion relative to glucose (**Figure 3G**), consistent with higher *SLC16A1* expression levels. These findings suggest that *in vitro* differentiation does not fully repress *SLC16A1* in control cells, potentially masking disease-specific differences and motivating further investigation in a more mature *in vivo* context.

### Loss of pyruvate-stimulated insulin secretion in WT but not in variant grafts

Our previous single-cell RNA sequencing data demonstrated incomplete *SLC16A1* silencing in SC-islets *in vitro*, whereas implantation *in vivo* resulted in complete silencing to levels comparable to primary human islets^32^. To determine whether this silencing effect would occur with the variant, we implanted equal numbers of WT and Var SC-islets under the kidney capsule of immunocompromised NOD-SCID gamma (NSG) mice and monitored the grafts for three months. At three months, membranous MCT1 expression was observed in the beta-cells of Var grafts but not in WT (**Figure 4A**). Quantification of MCT1 signal within the E-cadherin-defined beta-cell membrane confirmed significantly higher MCT1 levels in Var grafts (**Figure 4B**). To assess graft functionality in response to glucose and pyruvate, *ex vivo* GSIS and PSIS assays were performed (**Figure 4C**). Contrasting to the equivalent *in vitro* assay at S7w7, the WT grafts showed loss of pyruvate responsiveness, however the Var grafts retained pyruvate-induced insulin secretion, consistent with sustained MCT1 expression (**Figure 4D**).

**Figure 4.**
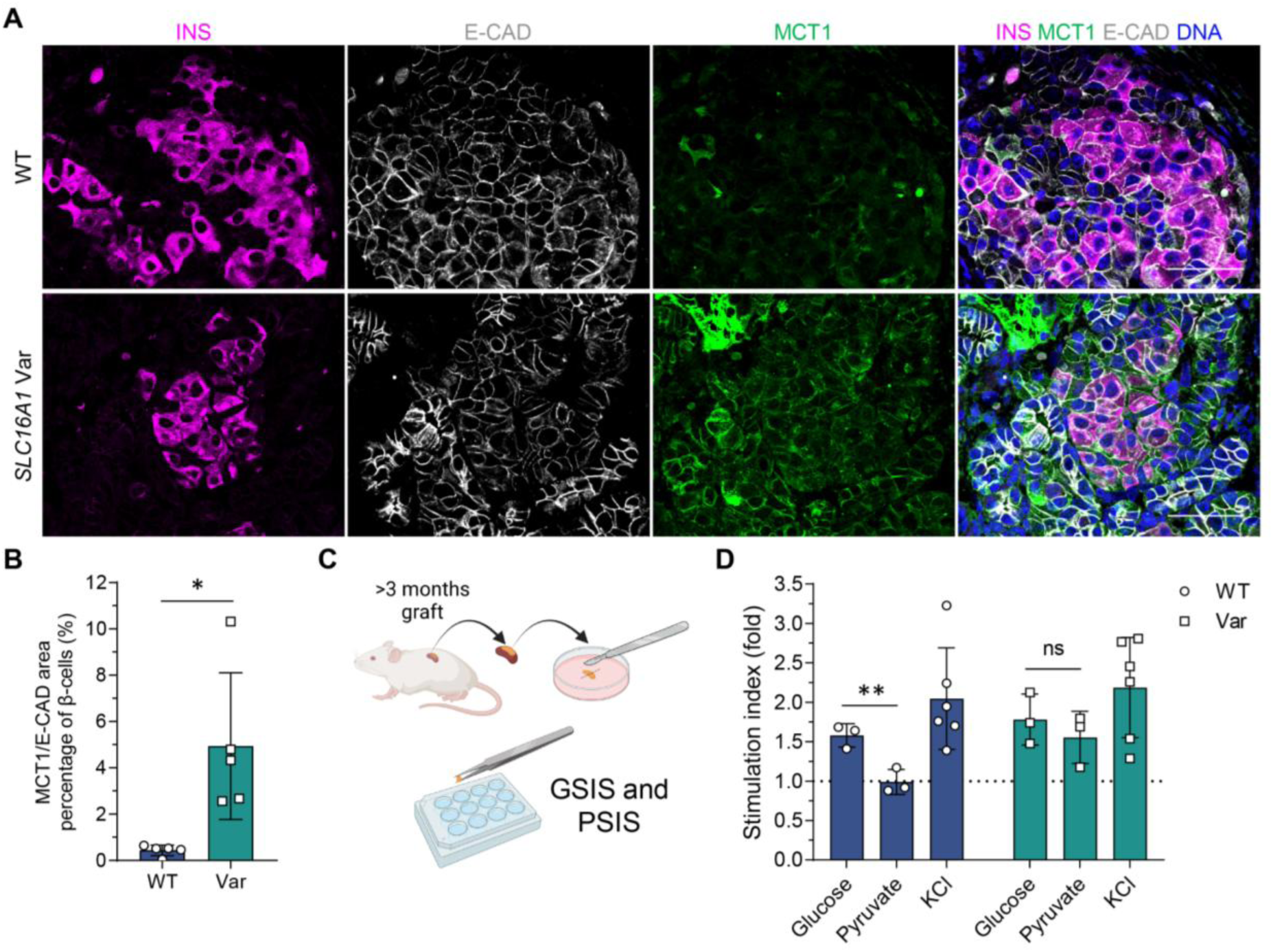
Elevated MCT1 expression in beta-cells in *SLC16A1* variant grafts. (A) Immunofluorescence images of INS⁺ E-CAD⁺ MCT1⁺ cells at 3-months post-implantation. Scale bar, 50 µm. (B) Quantification of the percentage of MCT1 signal area localised within the E-CAD-defined membrane region of INS⁺ cells (*n*=5). (C) Schematic representation of the *ex vivo* GSIS and PSIS assays after 3 months of implantation. Each implant was divided into two wells, each well for sequential GSIS or PSIS assay. (D) Insulin secretion stimulation index from static GSIS and PSIS *ex vivo* assays, shown as fold change relative to 2.8 mmol/L glucose. Conditions: Glucose (16.8 mmol/L), Pyruvate (0.5 mmol/L), and KCl (30 mmol/L) (*n*=3–6). Statistical significance in (B, D) was measured using unpaired *t*-test. Data are presented as means ±SD, \**p*<0.05, \*\**p*<0.01.

## Discussion

We identified a heterozygous pathogenic 94-bp deletion in the *SLC16A1* promoter in 37 individuals with childhood or adult-onset HI from 11 families. Genotyping identified a shared haplotype encompassing *SLC16A1* in eight individuals of Irish ancestry, consistent with a founder variant in this population. The identification of a *de novo* variant in one individual and the absence of the shared haplotype in one non-Irish case confirms that this is a recurrent variant that has arisen independently on multiple occasions. We hypothesize this is caused by a 17-bp motif which is repeated at the beginning and end of the deleted region.

Our immunohistochemistry data in pancreatic tissue from four individuals with the deletion provides conclusive evidence to support the hypothesis that *SLC16A1* promoter variants cause CHI through failed silencing of MCT1 in pancreatic beta-cells^10^. These findings were corroborated by functional studies using a patient-derived iPSCs model which demonstrated that variant SC-islets failed to repress MCT1 expression *in vitro* and *in vivo*. These studies further confirmed that in the variant cell line pyruvate uptake via MCT1 triggers inappropriate insulin secretion from beta-cells, explaining the clinical phenotype of exercise-induced HI. Together, these studies establish a direct causal link between this *SLC16A1* regulatory variant and dysregulated insulin secretion and offer a robust human-based platform for investigating this rare but clinically significant disorder.

The precise mechanism by which the 94-bp deletion and the other reported promoter variants in *SLC16A1* lead to aberrant expression of MCT1 in the pancreatic beta-cell is unknown and requires further study^10^. Chromatin accessibility data revealed a distinct peak overlapping the deleted region across multiple cell types, suggesting it is bound by as-yet unidentified regulatory factors. The beta-cell overall promoter accessibility is substantially lower than in other cell types that express *SLC16A1,* correlating with reduced gene expression, and suggesting that there is a beta-cell specific binding conferring repression at the promoter that is used in cells that express *SLC16A1*. This pattern contrasts with the mechanism proposed for *HK1*-HI where pathogenic variants disrupt a beta-cell-specific *cis*-regulatory element within an intron of the gene^16^.

All individuals with the 94-bp deletion had hypoglycemia with 62% receiving a formal diagnosis of HI. Notably, 16% of individuals reported a history of seizures suggesting significant neuroglycopenic symptoms. Consistently low HbA1c levels across the cohort also suggest that a lowering of glucose occurs in all patients, however the degree of reported symptoms was variable. In some cases, hypoglycemia had not been formally investigated or had not been recognised by the individual.

We found strong evidence that insulin-mediated hypoglycemia in *SLC16A1*-HI is not solely triggered by anaerobic exercise. While 19 individuals reported exercise-induced HI consistent with the earlier study^12^, 14 showed evidence of fasting-induced hypoglycemia, including through clinical fasting tests. There were also anecdotal reports of hypoglycemia following carbohydrate and protein-rich meals and episodes of emotional stress. Comprehensive clinical evaluation of affected individuals is needed to better characterise the full spectrum of triggers for inappropriate insulin secretion, which will improve our understanding of the underlying pathophysiology.

The approach to managing the hypoglycaemia differed between individuals. Three individuals continued to experience hypoglycemic symptoms despite undergoing partial pancreatectomy and 12 were receiving diazoxide treatment. Although most individuals (78%) were diagnosed in adulthood, the age at diagnosis ranged from 1.8 years to 78 years with many individuals reporting that symptoms worsened with age. Whilst some individuals may have adapted their lifestyles to reduce the frequency or severity of hypoglycaemic episodes, for example avoiding anaerobic exercise, eating frequent meals or limiting carbohydrate-rich foods and alcohol consumption, the low HbA1cs are in keeping with prolonged hypoglycemia in all individuals.

Identifying a monogenic cause of HI presenting beyond infancy is rare^33^. However, our discovery of pathogenic variants in multiple individuals who presented with HI for the first time in adulthood underscores the importance of considering genetic testing in all individuals with persistent HI of unknown cause, regardless of their age at diagnosis. This is particularly relevant in adults once more common causes of HI such as insulinoma have been excluded, or when a family history suggests a monogenic disorder. A genetic diagnosis of this condition has had a profound impact on many individuals in this study whose lives are significantly affected by a previously undiagnosed and difficult-to-manage disorder. For these individuals, these results mark the end of a lengthy and challenging diagnostic odyssey and represent the first tangible step towards improved understanding and management of their condition.

Given that pathogenic *SLC16A1* variants causing HI are non-coding, and that coding variants in this gene cause a distinct, unrelated disorder^34^, it is important to recognise the limitations of standard genetic testing methods when screening this gene. Firstly, although *SLC16A1* is often included on gene panels, routine sequencing typically targets protein-coding regions which would miss promoter variants unless these regions are specifically targeted. Consequently, such variants may go undetected, leading to missed diagnoses. Whilst this limitation is expected to diminish with the wider adoption of WGS, in the interim, laboratories offering HI genetic testing should ensure that the *SLC16A1* promoter region is adequately captured and analysed in all relevant referrals. A second consideration is whether the laboratory’s variant detection software can detect the 94-bp deletion. This medium-sized structural variant will be too large for some standard SNP/indel callers which typically detect variants up to 50-bp in size, but too small for structural variant callers which are built to detect variants >300-bp in size.

A further challenge faced by laboratories is in the interpretation of novel non-coding variants, especially in genes such as *SLC16A1* and *HK1*, where variable penetrance reduces the reliability of co-segregation analyses^17^. In this study, we provided strong evidence supporting the pathogenicity of these variants through a combination of clinical data, replication cohorts, analysis of patient pancreatic tissue, and patient iPS cell lines, resources that are not routinely available in most diagnostic laboratories but are necessary to accurately interpret the impact of novel variants on gene regulation.

In conclusion, we have identified a recurrent 94-bp deletion in the promoter region of the *SLC16A1* gene which encodes MCT1. We have shown that this deletion leads to the inappropriate expression of MCT1 in pancreatic beta-cells, resulting in HI due to the uptake of pyruvate which stimulates insulin secretion. These results enhance understanding of the pathophysiology of this disorder including the triggers for hypoglycemia which are not, as previously thought, limited to anaerobic exercise.

## Data Availability

Requests for collaboration and access to data will be considered by a steering committee following an application to the Genetic Beta Cell Research Bank (IRAS: 316,050, https://www.diabetesgenes.org/current-research/genetic-beta-cell-research-bank/).

## Acknowledgements

We are grateful to the families for participating in the study, particularly the index family who supported the writing of the clinical details section and Dr Marcia Bell who was involved in their clinical management. We thank Anna Ahmala and Nea Asumaa (Stem Cells and Metabolism Research Program, University of Helsinki) for their technical assistance and Dr Julieanne Knupp (University of Exeter) for providing bioinformatic support. We grateful to Prof Sarah Richardson for providing access to control pancreatic tissue stored in the Exeter Archival Diabetes Biobank. All IHC images were acquired and analysed in the RILD Bioimaging Facility, Exeter, UK with technical guidance from James Crichton. JMEM is the recipient of a European Society for Paediatric Endocrinology (ESPE) Research Fellowship and the Foundation for Paediatric Research Postdoctoral Fellowship, and NDLO is supported by a Wellcome Career Development Award (Grant Number: 227357/Z/23/Z). TWL is supported by the Academy of Medical Sciences/the Wellcome Trust/the Government Department of Science Innovation and Technology/the British Heart Foundation/Diabetes UK Springboard Award [SBF009\1135]. TO acknowledges the funding provided by the Academy of Finland (MetaStem Center of Excellence grant 312437), the Novo Nordisk Foundation, the Sigrid Juselius Foundation and the Wellcome Collaborative Award in Science. HI acknowledges the funding provided by the Finnish-Norwegian Medical Foundation and Orion Research Foundation. SEF has a Wellcome Trust Senior Research Fellowship (Grant Number 223187/Z/21/Z). MBJ is a Diabetes UK and Breakthrough T1D RD Lawrence Fellow (23/0006516). The clinical analysis and the study of human genetic and pancreatic tissue was supported by the National Institute for Health and Care Research Exeter Biomedical Research Centre. The views expressed are those of the author(s) and not necessarily those of the NIHR or the Department of Health and Social Care. We are grateful to Congenital Hyperinsulinism International (a 501©3 organisation) who support the costs of genetic testing in Exeter through the Open Hyperinsulinism Genes Project. The study sponsor/funders were not involved in the design of the study; the collection, analysis, and interpretation of data; writing the report; and did not impose any restrictions regarding the publication of the report.

## Author Contributions

JJB, HI, JMEM, TO and SEF designed the study. MFM, CSM, ON, JO, KN, SML, EP, CC, RC, JG, DO, YR and LGR recruited the patients. JJB, CSM, JALH, MNW, MBJ, TWL, and SEF performed molecular genetic analysis and/or interpretation of the genetic data. NS reviewed pathological material. JJB, JMEM, ATH and SEF analysed the clinical data. HI, MT, JRH, JSV, SE, EV, HM, DB and TO conducted the iPSC and/or IHC experiments. MV and NDLO conducted the epigenomic analysis. JJB, HI, JM, JRH, MV, NDLO, TO and SEF prepared the draft manuscript. JJB, HI, and JMEM contributed equally to this work. First-authorship order reflects their primary roles: JJB led genetic interpretation and clinical analysis; HI led iPSC-based disease modelling and functional assays; and JMEM led clinical-genetic integration and patient data analysis. All authors read and approved the final manuscript.

## Data Availability

The raw sequencing data generated during the current study are not publicly available to preserve patient confidentiality. Variant call format (.vcf) files are available through collaboration to experienced teams working on approved studies examining the mechanisms, cause, diagnosis and treatment of diabetes and other beta cell disorders. Requests for collaboration will be considered by a steering committee following an application to the Genetic Beta Cell Research Bank (IRAS: 316,050, https://www.diabetesgenes.org/current-research/genetic-beta-cell-research-bank/). Contact by email should be directed to Sarah Flanagan.

