## Supplementary Data for "A 94-bp Deletion in the Promoter of the Beta-Cell Disallowed gene *SLC16A1* causing Adult-onset Hyperinsulinism"

**Supplementary Information**

**Supplementary Methods**

*Targeted next generation sequencing (tNGS)*

In Exeter target enrichment of the *SLC16A1* promoter region was carried out using the Agilent SureSelect All Exon v5 capture kit (Agilent, Santa Clara, California). 100-bp paired-end reads were sequenced on a HiSeq 2500 (Illumina, San Diego, California). In Paris, target enrichment was performed using a custom Agilent tNGS panel (DIACHI_V7), sequencing was performed on a MiSeq (Illumina, San Diego, California) with sequence analysis performed using SeqNext V5.1. Analysis was conducted following GATK best practice guidelines, and sequence reads were aligned to the GRCh37 human reference genome using BWA-MEM.

*Immunofluorescence and quantification of MCT1 in patient beta-cells*

After dewaxing and rehydration, pancreatic samples from cases were subjected to heat-induced epitope retrieval (HIER) in 10 mM Tris Base, 1 mM EDTA, pH9 buffer for 20 min. After blocking for 10 mins with 5% normal goat serum (NGS) in antibody diluent / block (Akoya), the sections were probed in a sequential manner with rabbit monoclonal anti-MCT1 (Proteintech, 20139- 1-AP95452; 1/350, overnight at 4 degrees, then 1 hour at room temperature), followed by incubation with Opal Anti-mouse & Rabbit HRP secondary, then incubated with Opal fluorophore 520 (1/100, 10 min). Following washes in TBST, sections were then subjected to HIER for 6 mins on high, followed by 14 mins on simmer in 10 mM Citrate buffer (pH6). Following blocking as previously described, sections were probed with mouse monoclonal anti-insulin antibody (Affymetrix eBioscience, 14-9769-82, 1/800, 30 min), followed by incubation with Opal Anti-mouse & Rabbit HRP secondary, then incubated with Opal fluorophore 690, (1/100, 10 min). Next, following washes, sections were subjected to HIER for 6 mins on high, followed by 14 mins on simmer in 10 mM Citrate buffer (pH6). Following blocking as previously, cell nuclei were probed with DAPI (1/1000, 1hr). Sections were mounted using ProLong Diamond Antifade Mountant (Invitrogen, P36970). Once slides were dry, they were imaged at x40 using the PhenoImager HT 2.0 whole slide scanner (Akoya Bioscience) and then analysed in QuPath (V 0.4.3). The distribution of MCT1 and insulin were examined across the whole section, and the MFI of membrane MCT1 was plotted in each beta-cell. For higher resolution images and to determine cellular localisation, slides were imaged using a Leica DMi8 confocal microscope (Leica Microsystems).

*Generation of a patient-derived induced pluripotent stem cell model*

Dermal fibroblasts were obtained from a skin biopsy of an adult male with a heterozygous 94-bp deletion in the *SLC16A1* promoter. The fibroblasts were reprogrammed using CRISPR-activator system targeting the promoters of *OCT4*, *SOX2*, *KLF4*, *MYC*, *LIN28A*, miR-302/367, and the EEA motif (embryo genome activation-enriched Alu-motif). Briefly, one million cells were electroporated with 2 µg dCas9-activator plasmid (CRISPRa; Addgene #69535) and 4 µg guide plasmids (Addgene #102902, #201677) using the Neon transfection system with settings of 1650 V, 30 ms, and 3 pulses. The cells were cultured on Matrigel-coated (Corning, no. 354277; USA) 10 cm plates with MEF media. MEF media was changed daily until day 3, after which hES media with 0.25 mmol/L sodium butyrate (Sigma, B5887-1G) was added on days 4 and 5 with a 1:1 ratio. When stem cells emerged at day 6, Essential 8 medium (E8; Thermo Fisher, A1517001) was used for culturing. Stem cell colonies were picked into a 24-well plate for further expansion and characterization.

*In-vitro pancreatic endocrine differentiation culture*

Two million cells/well were seeded on 6-well Matrigel-coated plates or 12 million cells/10 cm plate in E8 medium supplemented with 10 µmol/L Rho-Associated kinase inhibitor (ROCKi, Y-27632). The differentiation was started 24 h after seeding and proceeded through a 7-stage differentiation (stages 1–7 [S1–S7]: S1–S4 in adherent culture; S4 in AggreWell [Stemcell Technologies, no. 34421; Vancouver, Canada] and S5–S7 in suspension culture).

*Flow cytometry*

To quantify the percentage of definitive endoderm-positive cells at stage 1, flow cytometry for C-X-C motif chemokine receptor 4 was performed on one million dissociated cells using TrypLE (Thermo Fisher Scientific) for 3 min at 37°C. Cells were resuspended in cold 5% vol/vol FBS-containing PBS and the conjugated antibodies were added and incubated for 30 min at room temperature. For other antigens flow cytometry at stages 4 and 7, cells were dissociated with TrypLE for 8 min at 37°C and resuspended in cold 5% FBS-containing PBS. A total of one million cells were fixed and permeabilised using Cytofix/Cytoperm (554714, BD Biosciences; New Jersey, USA) as per manufacturer's instructions. Primary, conjugated or IgG isotype (negative control) antibodies were incubated with the cells overnight at 4°C in Perm/Wash buffer (554714, BD Biosciences) containing 4% vol/vol FBS and then secondary antibodies for 45 min at room temperature. The cells were then analysed using a FACSCalibur cytometer (BD Biosciences) with BD Cellquest Pro v4.0.2 (BD Biosciences) and FlowJo software v10 (BD Biosciences). The details of antibodies and their dilutions for flow cytometry are given in **Supplementary Table 2**.

*Static glucose- and pyruvate-stimulated insulin secretion*

Fifty aggregates were randomly picked and preincubated in 2.8 mmol/L glucose Krebs buffer in a 12-well plate placed on a rotating platform at 95 rpm/min for 90 min at 37°C. Aggregates were then washed with Krebs buffer and sequentially incubated in Krebs buffer containing 2.8 mmol/L glucose, 16.8 mmol/L glucose (PSIS; 2.8 mmol/L glucose plus 0.5 mmol/L sodium pyruvate) and 2.8 mmol/L glucose plus 30 mmol/L KCl, on the rotating platform for periods of 30 min each. Samples of 200 µl were collected from each treatment and stored at −80°C for insulin ELISA measurements (Mercodia; Uppsala, Sweden). Following the assay, the SC-islets were collected, and the total insulin and DNA contents were analysed. Insulin content results were normalised to the DNA content of the beta-cell fraction, calculated from flow cytometry insulin-positive cell percentage. For *ex vivo* assays three months post-engraftment, mice were euthanized, and graft-bearing kidneys were retrieved. The grafts were dissected using forceps from the inner surface of the peeled kidney capsule and cut into three portions: one for GSIS, one for PSIS, and one for immunohistochemistry (IHC). For secretion assays, graft pieces were incubated overnight in Stage 7 CMRL medium (1 ml per well in a 12-well plate). GSIS and PSIS assays were performed the following day, following the same protocol as for Stage 7 aggregates. After the assay, graft pieces were collected for DNA quantification and insulin content analysis.

*Immunocytochemistry and immunohistochemistry*

For adherent culture immunocytochemistry, cells were fixed with 4% wt/vol paraformaldehyde (PFA) for 15 min at room temperature, permeabilised with 0.5% vol/vol Triton X-100 in PBS for 15 min at room temperature, then blocked with Ultra V block (Thermo Fisher Scientific) for 10 min followed by incubation with primary antibodies overnight at 4°C and secondary antibodies for 1 h at room temperature diluted in 0.1% vol/vol Tween in PBS. For paraffin embedding, aggregates were fixed with 4% PFA overnight at 4°C then embedded in 2% low-melting agarose (Fisher Bioreagents; USA) PBS and transferred to paraffin blocks. Implanted grafts were retrieved, dissected and fixed with 4% PFA at room temperature for 48 h before being paraffin embedded and cut into 5 µm sections using Leica microtome. For immunohistochemistry, slides were deparaffinised and antigen-retrieved by boiling slides in 0.1 mol/L citrate buffer (pH 6) using a Decloaking chamber (Biocare Medical; USA) at 95°C for 20 min. For MCT1 antigen retrieving slides were boiled in Tris-EDTA Buffer: 10 mmol/L Tris Base, 1 mmol/L EDTA Solution (pH 9) using a Decloaking chamber at 95°C for 20 min. EVOS FL Digital Inverted Fluorescence Microscope (Invitrogen; USA) or a Zeiss Axio Observer Z1 with Apotome (ZEN-2 software; Germany) were used for image acquisition. All samples were randomised, blinded, equally treated and acquired with the same microscope parameters. MCT1, ECAD, and INS positive areas were quantified using CellProfiler 4.2.8. First, the insulin-positive area was masked, followed by quantification of ECAD within the insulin-positive mask. MCT1 signal colocalized with the masked ECAD area was then quantified and plotted. The details of antibodies and their dilutions used in the study for immunofluorescence are described in **Supplementary Table 2**.

*RNA extraction and RT-qPCR*

Total RNA was extracted using a NucleoSpin RNA Plus kit (Macherey-Nagel; Germany) for lysed samples of adherent cells or NucleoSpin RNA Plus XS kit (Macherey-Nagel; Germany) for samples of aggregates. A total of 500-1500 ng RNA was reversely transcribed using 0.5 µl Moloney murine leukaemia virus reverse transcriptase (Promega, M1701; USA), 4 µl RT buffer 5× (Promega), 2.5 µl dNTPs 2.5 mmol/L, 1 µl Oligo-dT 500 µg/ml (Promega), 0.2 µl Random hexamers 500 µg/ml (Promega) and 0.5 µl Riboblock RNase inhibitor 40 u/µl (Fermentas; USA) for 90 min at 37°C. The generated cDNA was amplified using 5× HOT FIREPol EvaGreen qPCR Mix Plus no ROX (Solisbiodyne; Estonia) in a 20 µl reaction. The reactions were pipetted using QIAgility (Qiagen; Germany) robot into a 100-well disc run in Rotor-Gene Q. Relative quantification of gene expression was analysed using the method, with cyclophilin G (PPIG) as a reference gene. Reverse transcription without template was used as a negative control and an exogenous positive control was used as a batch calibrator. The RT-qPCR primers sequences are described in **Supplementary Table 3.**

*Quantification and statistical analysis.*

Data were collected from at least three independent differentiation experiments. Blinding was applied for immunohistochemical quantification. Morphological data represent population-wide observation from independent differentiation experiments. All representative images in the figures were reproducible in all the experiments performed and are followed by a quantification panel for all the experiments; the n value represents independent differentiation experiments, unless otherwise stated. Bar graphs are represented showing all the individual data points. Statistical methods used are described in each figure legend and individual method section. The results are presented as the mean ± SD unless otherwise stated. p values<0.05 were considered statistically significant.

**Animal care experiments**

Animal care and experiments were approved by the National Animal Experiment Board in Finland (ESAVI/14852/2018). NOD.Cg-Prkdcscid Il2rgtm1Wjl/SzJ (NOD-SCID-Gamma [NSG], 005557, Jackson Laboratory; USA,) mice were obtained from SCANBUR and housed at Biomedicum Helsinki animal facility, on a 12 h light–dark cycle and fed standard chow ad libitum (2016 Teklad global 16% protein rodent diet, ENVIGO; USA). The temperature was kept at 23°C with 24% relative humidity.

### Supplementary Results

*Characterization of patient iPSCs and quality control of differentiation into pancreatic endocrine cells.*

Patient-specific iPSCs were derived from fibroblasts. Two iPSC clones (Var1 and Var2) from the same reprogramming experiment were validated and were found to express pluripotency markers and *SLC16A1* at similar levels to control WT iPSCs (**Supplementary Figure 6A, B**). Patient and control iPSCs were differentiated in parallel into pancreatic endocrine cells using a seven-stage protocol to generate stem-cell-derived islets (SC-islets). Differentiation efficiency was similar between groups, with comparable proportions of $CXCR4^+$ definitive endoderm cells at stage 1 (S1) (**Supplementary Figure 6C, D**), and similar populations of $PDX1^+$/$NKX6.1^+$ pancreatic progenitors, $SOX9^+$ cells, $NEUROG3^+$ endocrine progenitors, and $CHGA^+$ endocrine cells at stage 4 (S4) (**Supplementary Figure 6E–H**).

By week 3 of the final stage (S7w3), Var SC-islets showed comparable $PDX1^+$/$NKX6.1^+$ populations to WT, with a modest reduction in $CHGA^+$ cells (94.7 ± 1.7% WT vs. 84 ± 3.4% Var) (**Supplementary Figure 7A–E**). Glucagon-positive ($GCG^+$) alpha cells were similar between groups, whereas insulin-positive ($INS^+$) beta-cells were reduced in Var SC-islets (**Supplementary Figure 7F–H**). These differences likely reflect donor-specific genetic background rather than the *SLC16A1* variant itself, as *SLC16A1* expression remained unchanged until 7 weeks after endocrine cell formation. Overall, both Var and WT lines produced sufficient $INS^+$ cells to allow assessment of beta-cell functionality at S7w7 (**Figure 3F**).

**Supplementary Tables**

**Supplementary Table 1:** Details of PCR primer sequences and methodology for Sanger sequencing.

| Primer direction | Sequence (5'-3') | Region amplified - GRCh38 | Sequencer | Analysis software |
|---|---|---|---|---|
| Forward | AGGCCTCTCGGTGACTTTTC | 1:112955847-112956580 (734-bp) | ABI 3730 capillary machine (Applied Biosystems) | Mutation Surveyor version 3.24 software (SoftGenetics) |
| Reverse | AAGGTCTCCTTCACCAGCAC | | | |
| Forward | GATTGCCTAGAGCTCGTCAGA | 1:112956016-112956502 (487-bp) | | SeqScape (Thermo Fisher Scientific, Waltham, Massachusetts, USA) |
| Reverse | CGGCTGTTACCCAACTAACC | | | |
| Forward | CGGGTCGGACTGGGACG | 1:112956035-112956415 (381 bp) | | Geneious Prime |
| Reverse | CGTTATATGCGCGGATCGCAG | | | |

**Supplementary Table 2:** Antibodies used for immunofluorescence and flow cytometry.

| Antibody for ICC and IHC | Manufacturer | Catalog # | Dilution |
|---|---|---|---|
| Donkey anti-Goat IgG secondary, Alexa Fluor™ 488 | Thermo Fisher Scientific | A-11055 | 1:500 |
| Donkey anti-Mouse IgG secondary, Alexa Fluor™ 488 | Thermo Fisher Scientific | A-21202 | 1:500 |
| Donkey anti-Mouse IgG secondary, Alexa Fluor™ 594 | Thermo Fisher Scientific | A-21203 | 1:500 |
| Donkey anti-Rabbit IgG secondary, Alexa Fluor™ 488 | Thermo Fisher Scientific | A-21206 | 1:500 |
| Donkey anti-Sheep IgG secondary, Alexa Fluor™ 594 | Thermo Fisher Scientific | A- 11016 | 1:500 |
| FLEX Polyclonal Guinea Pig Anti-Insulin, Ready-to-Use | Agilent | IR002 | 1:2 |
| Goat anti-Guinea Pig IgG secondary, Alexa Fluor™ 594 | Thermo Fisher Scientific | A-11076 | 1:500 |
| Goat anti-Guinea Pig IgG secondary, Alexa Fluor™ 633 | Thermo Fisher Scientific | A-21105 | 1:500 |
| Goat anti-Mouse IgG (H+L) Cross-Adsorbed Secondary Antibody, Alexa Fluor™ 555 | Thermo Fisher Scientific | A21422 | 1:500 |
| Goat anti-PDX1 | R&D systems | AF2419 | 1:250 |
| Goat anti-Rabbit IgG (H+L) Cross-Adsorbed Secondary Antibody, Alexa Fluor™ 647 | Thermo Fisher Scientific | A21244 | 1:500 |
| Monoclonal mouse anti-E-Cadherin | BD Biosciences | 610181 | 1:250 |
| Monoclonal mouse anti-Glucagon | Sigma-aldrich | G2654 | 1:300 |
| Monoclonal mouse anti-Human Chromogranin A | Dako | M0869 | 1:500 |
| Mouse anti-CHGA | Dako | M0869 | 1:500 |
| Mouse anti-GCG antibody | Sigma-Aldrich | G2654 | 1:500 |
| Mouse anti-Nanog mAb | Thermo Fisher Scientific | MA1-017 | 1:100 |
| Mouse anti-NKX6.1 | DSHB Hybridoma | F55A10 | 1:250 |
| Mouse anti-OCT4 mAb | Thermo Fisher Scientific | MA1-104 | 1:100 |
| Polyclonal rabbit anti-MCT1 | Proteintech | 20139-1-AP | 1:250 |
| Rabbit anti-SOX9 | Sigma-Aldrich | AB5535 | 1:500 |
| Sheep anti Human Neurogenin-3 | R&D systems | AF3444 | 1:250 |
| **Antibody for Flow cytometry** | **Manufacturer** | **Catalog #** | **Dilution** |
| Alexa Fluor 647 Mouse anti NKX6-1 | BD Biosciences | 563338 | 1:80 |
| Alexa Fluor 647 Mouse IgG1 k isotype control | BD Biosciences | 557714 | 1:80 |
| Alexa Fluor 647 Rabbit anti Insulin (C27C9) | Cell Signaling Technology | 9008 | 1:80 |
| Alexa Fluor 647 Rabbit IgG Isotype Control | Cell Signaling Technology | 3452S | 1:80 |
| Monoclonal mouse anti-Glucagon | Sigma-aldrich | G2654 | 1:160 |
| Monoclonal mouse anti-Human Chromogranin A | Dako | M0869 | 1:80 |
| Mouse Anti-CD184 (CXCR4) Monoclonal Antibody | BD Biosciences | 555974 | 1:10 |
| Mouse IgG2a, kappa Isotype Control, Phycoerythrin Conjugated | BD Biosciences | 5555749 | 1:10 |
| PE-Mouse anti PDX1 | BD Biosciences | 562161 | 1:80 |

**Supplementary Table 3:** RT-qPCR primers used for iPSCs.

| Gene | Forward Primer (5’-3’) | Reverse Primer (5’-3’) | NCBI reference | Amplicon size (bp) |
|---|---|---|---|---|
| *PPIG* | TCTTGTCAATGGCCAACAGAG | GCCCATCTAAATGAGGAGTTG | NM_004792 | 84 |
| *NANOG* | CTCAGCCTCCAGCAGATGC | TAGATTTCATTCTCTGGTTCTGG | NM_024865.2 | 94 |
| *OCT4* | TTGGGCTCGAGAAGGATGTG | TCCTCTCGTTGTGCATAGTCG | NM_002701 | 91 |
| *SLC16A1* | GTTGGACCCCAGAGGTTCTC | TTGAGCCGACCTAAAAGTGG | NM_003051.4 | 95 |
| *SOX2* | GCCCTGCAGTACAACTCCAT | TGCCCTGCTGCGAGTAGGA | NM_003106 | 85 |

**Supplementary Table 4:** List of 23 novel heterozygous non-coding variants intersecting with a beta-cell accessible region, shared between all five members of the index family.

| Variant (GRCh38) | Closest gene |
|---|---|
| 1:112619168 G>A | *ST7L* |
| 1:112,956,279 CCGCCCCCTCCCCGCCACACAGACATCCGAACTGCAGCCCGCGCCTCCA CACGCTTTCAGCCGCGCGCGCCCTCTAGCTCGCCCGCGCGCGCCGG>C | *SLC16A1* |
| 1:115469795 T>C | *NGF* |
| 1:116138981 A>G | *MAB21L3* |
| 1:150213526 G>A | *ANP32E* |
| 1:150363457 G>C | *RPRD2* |
| 2:170123454 T>C | *UBR3* |
| 2:170156923 T>C | *MYO3B* |
| 2:170324899 A>C | *MYO3B* |
| 2:177553066 C>T | *AGPS* |
| 2:178342182-178342183 delinsAA | *OSBPL6* |
| 2:178523603 G>T | *TTN-AS1* |
| 2:184597921 C>T | *ZNF804A* |
| 2:186422491-186422493delinsAAGCATGTCACATGGCATG | *LINC01473* |
| 2:191311865 G>A | *MYO1B* |
| 2:205347744 G>T | *PARD3B* |
| 2:207258513 T>G | *MYOSLID* |
| 2:209615138 A>G | *MAP2* |
| 2:214569727 C>A | *VWC2L* |
| 2:215836217 C>T | *LINC00607* |
| 2:215920574 C>T | *LINC00607* |
| 2:217511077 C>T | *DIRC3* |
| 21:30228051 G>A | *CLDN8* |

**Supplementary Table 5:** Variant classification details for the novel 94-bp *SLC16A1* promoter deletion.

| Criteria used | Explanation |
|---|---|
| PS2_strong | *De novo* (maternity and paternity confirmed) in one case |
| PP1_strong | Co-segregation with disease in multiple affected family members in a gene definitively known to cause the disease(1), in pedigree 2. |
| PS4_moderate | Prevalence significantly increased in cases over controls. At least 3 separate occurrences of the variant (Irish founder variant, non-Irish case without the haplotype and *de novo* in one case) |

The variant was classified as pathogenic according to the ACGS, ACMG and non-coding variant guidelines.(2-4)

**Supplementary Table 6:** Example of data from a formal fasting test performed on one case with the *SLC16A1* 94-bp promoter deletion.

| **Time (hours)** | **Glucose (mmol/L)** | **Insulin (pmol/L)** |
|---|---|---|
| 0 | 4.2 | 76.4 |
| 1 | 3.2 | 97.2 |
| 1.5 | 3.1 | 76.4 |
| 2 | 3.1 | 69.4 |
| 2.5 | 2.9 | 34.7 |
| 3 | 3.1 | 27.8 |
| 3.5 | 3.4 | 97.2 |
| 4 | 2.6 | 62.5 |
| 4.5 | 2.2 | 27.8 |
| 5 | 2.8 | 27.8 |

Grey shading indicates time points in which the individual had hypoglycemia with blood glucose <2.8 mmol/L.

### Supplementary Figures

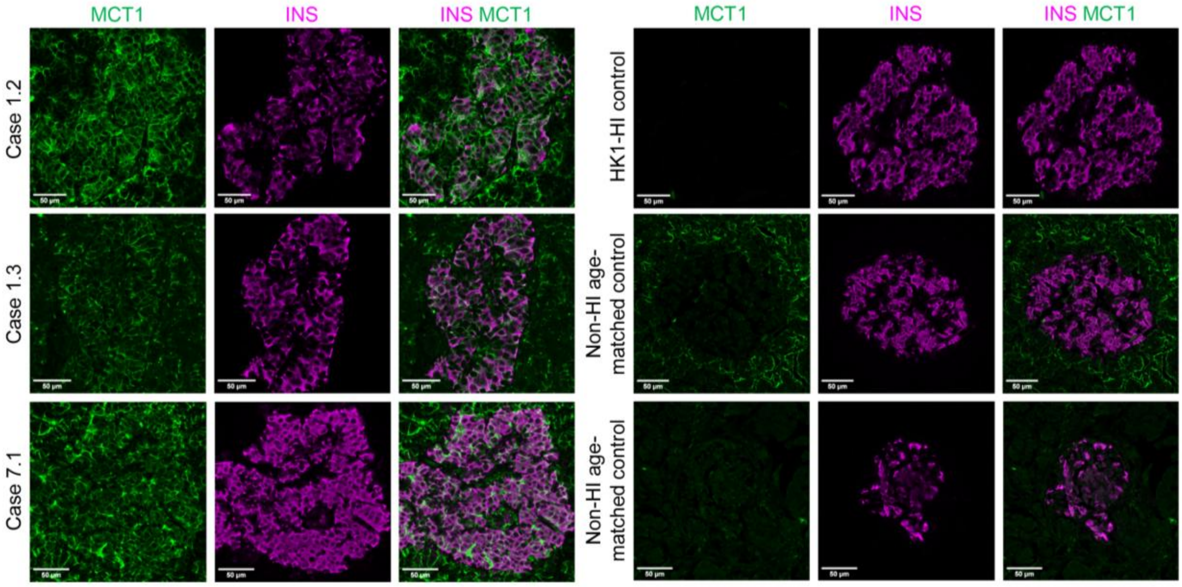


**Supplementary Figure 1. MCT1 is present in the beta-cell membrane of individuals with the 94-bp promoter deletion but not in the controls.**
Immunohistochemical staining of MCT1 (green) and insulin (magenta) was performed on pancreatic tissue resected from four patients all of whom are heterozygous for a 94-bp deletion in the *SLC16A1* promoter. Imaging shows that MCT1 localizes to the beta-cell membrane of case islets, as indicated by distinct membranous staining pattern and localisation with insulin staining, and to the cells of the surrounding exocrine tissue. In contrast, no membranous expression of MCT1 is seen in the beta-cells of the *HK1*-HI control or the age-matched controls and only minimal membranous expression is seen in a subset of the exocrine cells.

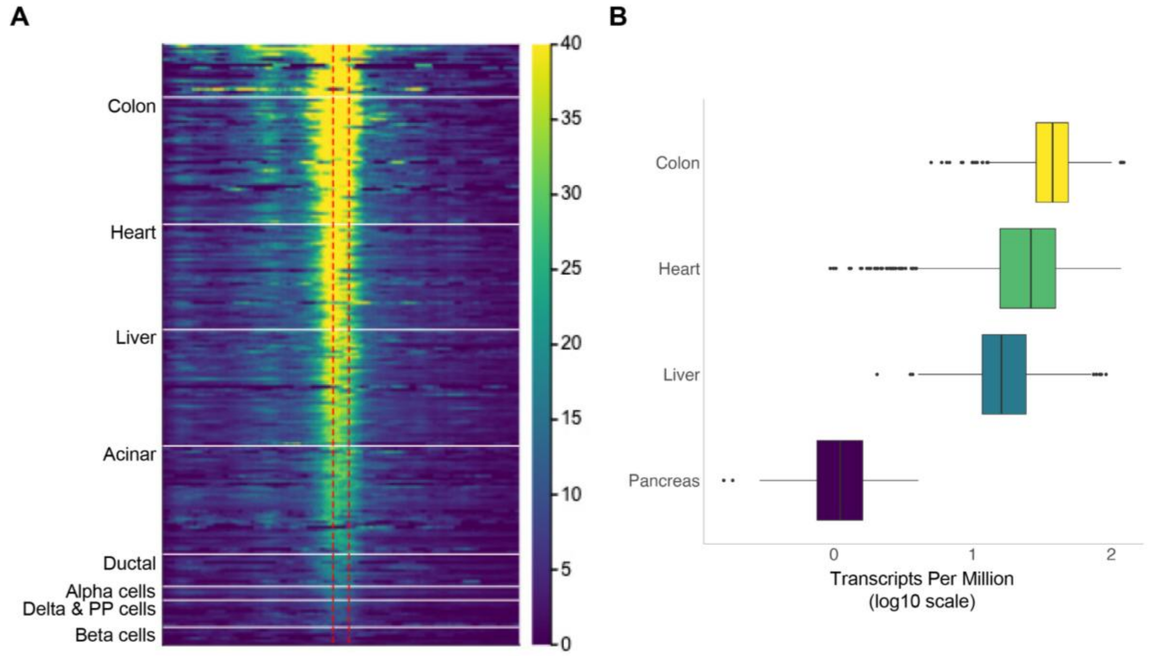


**Supplementary Figure 2. Accessibility of the *SLC16A1* 94-bp promoter deletion region across 222 cell types.**

(**A**) Heatmap showing chromatin accessibility at *SLC16A1* promoter, 94-bp promoter deletion region, indicated by red vertical dashed lines, and 1 kb either side, across 222 cell types ordered by maximal chromatin accessibility. Pancreatic cell types and cell types with higher *SLC16A1* expression than pancreas highlighted.

(**B**) Boxplots depicting gene level *SLC16A1* expression (Transcripts per Million) across pancreas and five tissues with higher *SLC16A1* expression compared to pancreas, from the version 8 release of the Genotype-Tissue Expression GTEx project. Tissues are ordered by median expression, with lowest expression in pancreas. Each boxplot central line denotes median, box limit the interquartile range, and whiskers extend to furthest point within 1.5 interquartile range, with points outside this range shown as outliers exceeding whiskers (colon; colon-sigmoid and colon-transverse, heart; heart- left ventricle and heart- atrial appendage). Although, beta-cells have a clear peak of chromatin accessibility (Figure 2C), overall promoter accessibility is substantially reduced compared to cell types that express *SLC16A1* and proportional to gene expression.

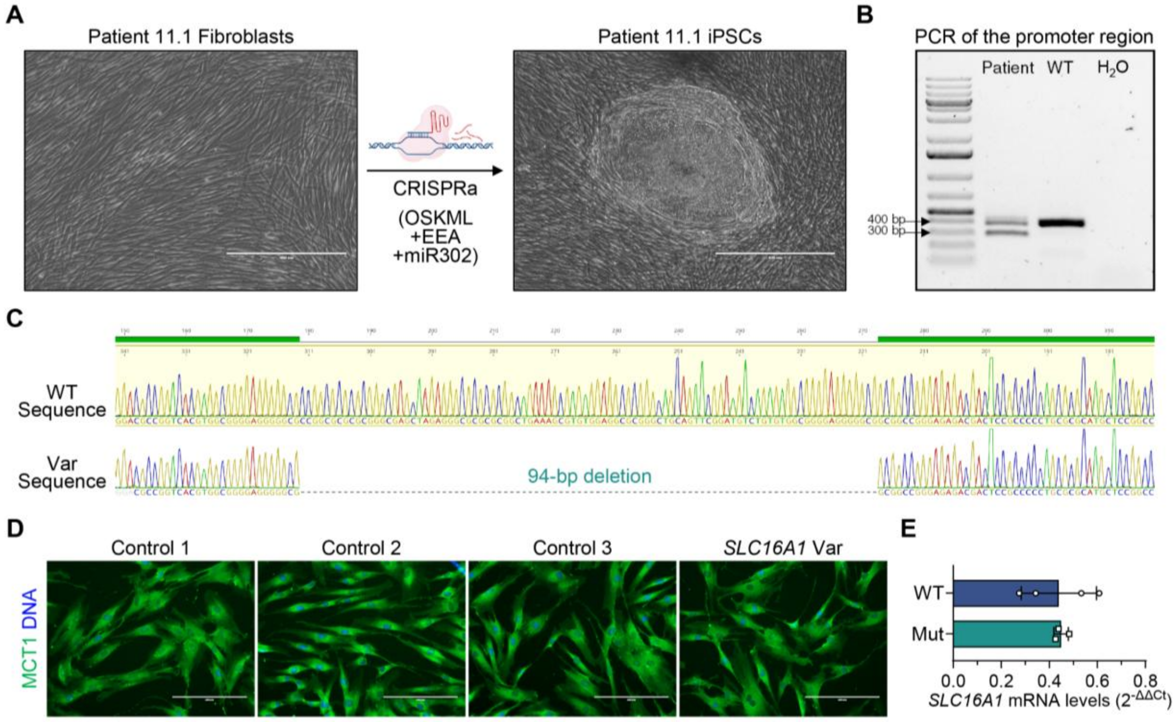


**Supplementary Figure 3. Generation of iPSCs from patient skin fibroblasts and characterization of MCT1 expression in fibroblasts.**

(**A**) Phase contrast images of patient fibroblasts before and after electroporation with the CRISPRa reprogramming system. The CRISPRa system targets *OCT4*, *SOX2*, *KLF4*, *MYC*, and *LIN28A* (OSKML), along with an EEA motif (embryo genome activation-enriched Alu-motif) and the miRNA miR-302. The right panel shows one of the stem cell colonies at day 18 post-electroporation. Scale bar, 400 µm. CRISPRa illustration was created with BioRender.com.

(**B**) PCR gel electrophoresis illustrating the heterozygous deletion in the patient sample.

(**C**) Sanger sequencing of the smaller PCR band—gel-purified from the heterozygous patient sample—displaying the 94-bp deletion. Sequence alignment and illustration was created using Geneious Prime.

(**D**) Immunofluorescence images of fibroblasts showing comparable MCT1 expression between the patient sample (*SLC16A1* Var) and different control donors. Scale bar, 200 µm.

(**E**) Relative gene expression levels of *SLC16A1* in the samples shown in (D) (*n*=3–4). Statistical significance was measured using unpaired Mann-Whitney test. Data are presented as means ±SD.

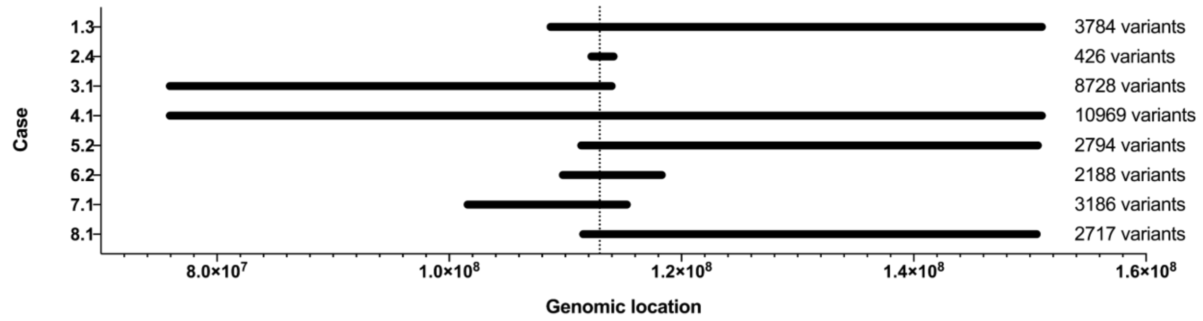


**Supplementary Figure 4. Schematic representation of the haplotype analysis results.** The genomic location on chromosome 1 along the x-axis. Each bar represents the region of shared genetic variants from the SNP array data for each family with the shared haplotype. The location of the *SLC16A1* promoter variant (GRCh38:112,956,297-112,956,390del) is indicated by the dashed line. The number of consecutive shared variants for each individual is provided to the right of the figure.

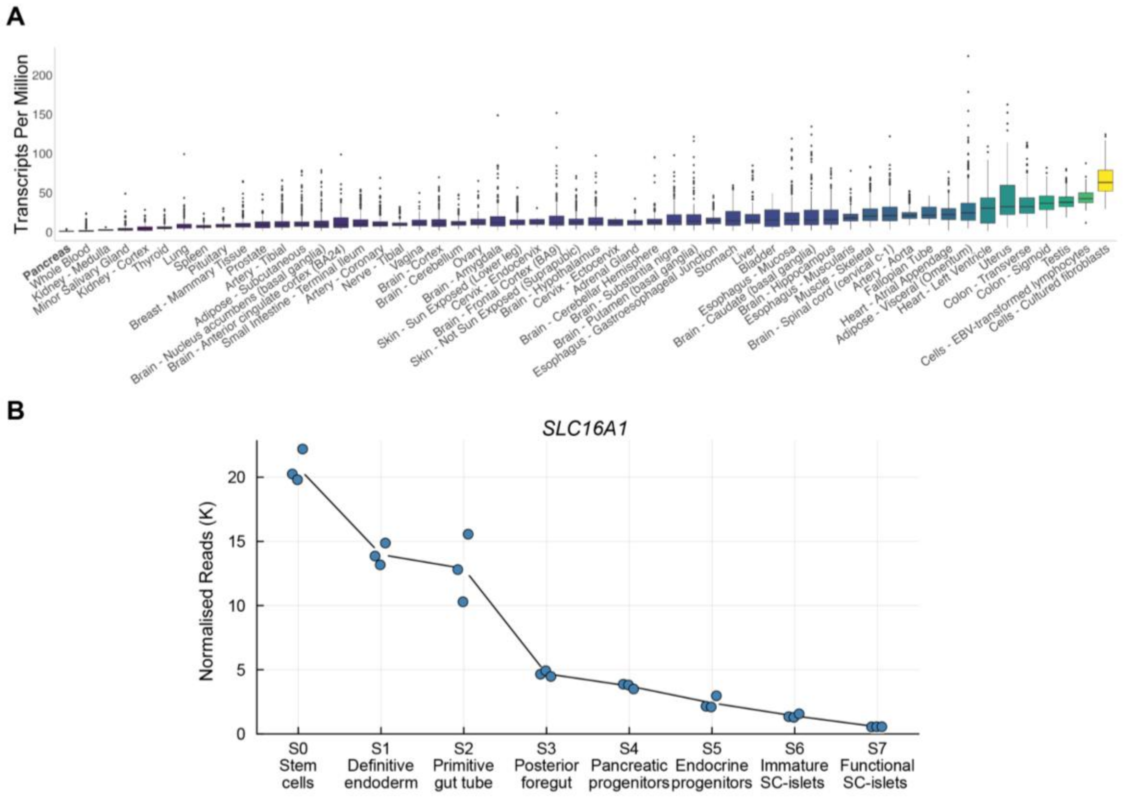


**Supplementary Figure 5. *SLC16A1* expression across 54 tissues in GTEx and across stem-cell pancreatic differentiation.**

(**A**) Boxplots describing gene level *SLC16A1* expression (Transcripts per Million) across 54 tissues, from the version 8 release of the Genotype-Tissue Expression GTEx project (5). Tissues are ordered by median expression, with lowest expression in pancreas (in bold). Each boxplot central line denotes median, box limit the interquartile range, and whiskers extend to furthest point within 1.5 interquartile range, with points outside this range shown as outliers exceeding whiskers.

(**B**) Gene expression levels of *SLC16A1* as normalized read counts (in thousands) across the 7-stage differentiation protocol of hESC H1 cells toward stem-cell-derived islets (SC-islets), based on reanalysis of bulk RNA sequencing data from De Franco *et al.* (*n*=3) (6). Data are shown as mean values with individual datapoints.

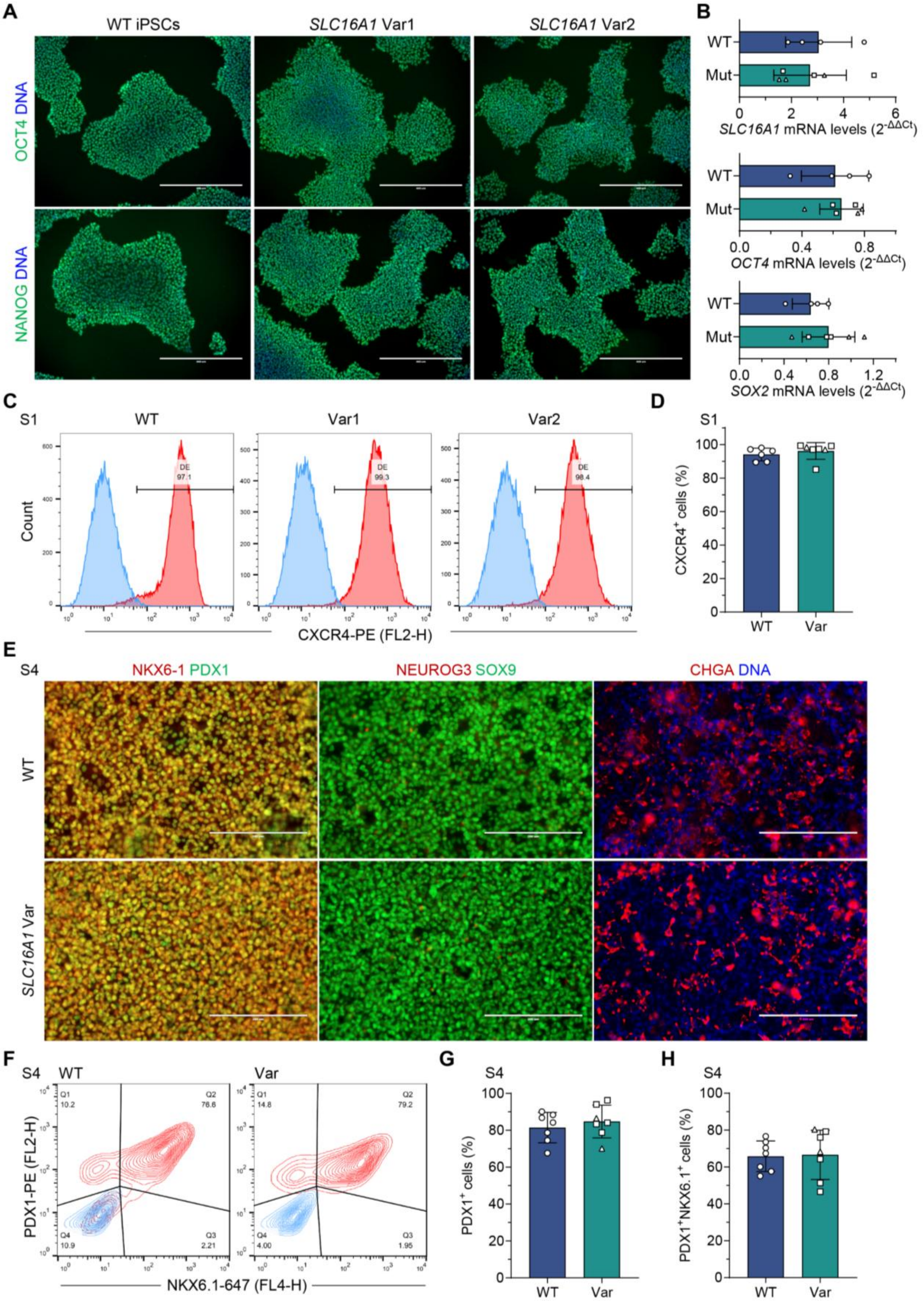
A
WT iPSCs
SLC16A1 Var1
SLC16A1 Var2
OCT4 DNA
NANOG DNA
B
WT
Mut
SLC16A1 mRNA levels (2^-ΔΔCt)
OCT4 mRNA levels (2^-ΔΔCt)
SOX2 mRNA levels (2^-ΔΔCt)
C
S1
WT
Var1
Var2
Count
DE
97.1
99.3
98.4
CXCR4-PE (FL2-H)
D
S1
CXCR4+ cells (%)
WT
Var
E
S4
NKX6-1 PDX1
NEUROG3 SOX9
CHGA DNA
WT
SLC16A1 Var
F
S4
WT
Var
Q1
10.2
Q2
78.6
Q4
10.9
Q3
2.21
Q1
14.8
Q2
79.2
Q4
4.00
Q3
1.95
PDX1-PE (FL2-H)
NKX6.1-647 (FL4-H)
G
S4
PDX1+ cells (%)
WT
Var
H
S4
PDX1+NKX6.1+ cells (%)
WT
Var

**Supplementary Figure 6. Characterisation of patient-derived iPSCs and assessment of the *SLC16A1* promoter variant on their differentiation into pancreatic progenitors.**
(**A**) Immunofluorescence images of iPSCs demonstrating similar OCT4 and NANOG expression between the patient samples (*SLC16A1* Var1 and *SLC16A1* Var2) and the healthy control iPSC-line HEL24.3 (WT). Scale bar, 400 µm.
(**B**) Relative gene expression levels of *SLC16A1, OCT4* and *SOX2* among the samples shown in (A), with *SLC16A1* Var1 represented by square datapoints and *SLC16A1* Var2 represented by triangle datapoints ($n$=4–6).

(**C**) Flow cytometry analysis of C-X-C motif chemokine receptor 4-positive ($CXCR4^+$) cells at the end of the definitive endoderm stage S1. CXCR4 sample is shown in red and IgG isotype negative control is shown in blue.
(**D**) Quantification of (C) ($n$=5–7).

(**E**) Immunofluorescence images showing $PDX1^+$ and $NKX6.1^+$, $NEUROG3^+$ and $SOX9^+$, and $CHGA^+$ cells at the end of the pancreatic progenitor stage S4. Scale bar, 200 µm.

(**F**) Flow cytometry analysis of $PDX1^+$ and $NKX6.1^+$ cells at S4. PDX1 and NKX6.1 sample is shown in red and IgG isotype negative control is shown in blue.

(**G**) $PDX1^+$ cell percentage quantified from (F) ($n$=7).

(**H**) $NKX6.1^+PDX1^+$ cell percentage quantified from (F) ($n$=7).
*SLC16A1* Var1 represented by square datapoints and *SLC16A1* Var2 represented by triangle datapoints. Statistical significance in (B, D, G and H) was measured using unpaired Mann-Whitney test. Data are presented as means ±SD.

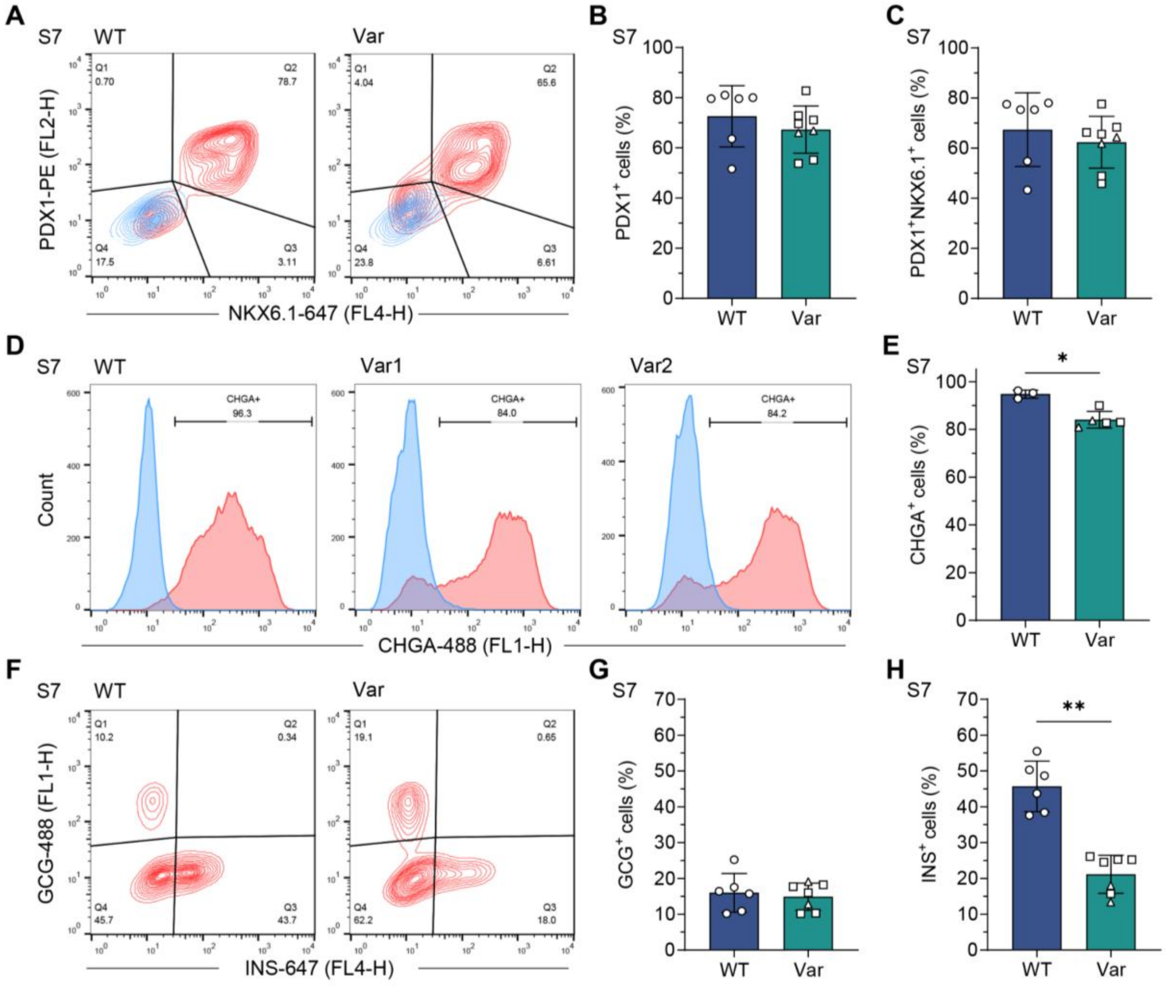


**Supplementary Figure 7. Characterisation of cell type composition at stage 7 of SC-islets.**

(**A**) Flow cytometry analysis of PDX1$^+$ and NKX6.1$^+$ cells at S7w3. Sample is shown in red and IgG isotype negative control is shown in blue.

(**B**) PDX1$^+$ cell percentage quantified from (A) (*n*= 6–8).

(**C**) NKX6.1$^+$PDX1$^+$ cell percentage quantified from (A) (*n*=6–8).

(**D**) Flow cytometry analysis of CHGA$^+$ cells at S7w3. Sample is shown in red and secondary antibody-treated negative control is shown in blue.

(**E**) CHGA$^+$ cell percentage quantified from (D) (*n*=3–5).

(**F**) Flow cytometry analysis of GCG$^+$ and INS$^+$ cells at S7w3.

(**G**) GCG$^+$ cell percentage quantified from (F) (*n*=6–7).

(**H**) INS$^+$ cell percentage quantified from (F) (*n*=6–7).

Statistical significance in (A–H) was measured using unpaired Mann-Whitney test. Data are presented as means ±SD, *$p$<0.05, **$p$<0.01.

**Acknowledgements**

The Genotype-Tissue Expression (GTEx) Project was supported by the Common Fund of the Office of the Director of the National Institutes of Health, and by NCI, NHGRI, NHLBI, NIDA, NIMH, and NINDS. The data used for the analyses described in this manuscript were obtained from the GTEx Portal on 10/14/2024.
